# Data-driven assessment of adolescents’ mental health during the COVID-19 pandemic

**DOI:** 10.1101/2022.01.06.22268809

**Authors:** Yonatan Bilu, Natalie Flaks-Manov, Maytal Bivas-Benita, Pinchas Akiva, Nir Kalkstein, Yoav Yehezkelli, Miri Mizrahi-Reuveni, Anat Ekka-Zohar, Shirley Shapiro Ben David, Uri Lerner, Gilad Bodenheimer, Shira Greenfeld

## Abstract

**Importance:** Adolescents’ mental health and well-being were severely compromised during the COVID-19 pandemic. Longitudinal follow-up studies, based on real-world data, assessing the changes in mental health of adolescents during the later phase of the COVID-19 pandemic are needed.

**Objective:** To quantify the effect of COVID-19 on the incidence of Israeli adolescents’ mental health outcomes from electronic health record (EHR) data.

**Design, Setting and Participants:** Retrospective cohort study analyzing EHR data of Maccabi Healthcare Services members, the second largest Health Maintenance Organization in Israel. Eligible subjects were 12-17 years old, during 2017-2021 with no previous diagnosis or psychiatric drug dispensation of those analyzed in this study. This resulted in over 200,000 eligible participants each year.

**Exposure:** COVID-19 pandemic and the measures taken to mitigate it.

**Main Outcomes and Measures:** Incidence rates of mental health diagnoses (depression; anxiety; obsessive-compulsive disorder; stress; eating disorders; ADHD), and psychiatric drugs dispensation (antidepressants; anxiolytics; antipsychotics; ADHD agents) were measured, and relative risks were computed between the years. Subgroup analyses were performed for age, gender, population sector and socioeconomic status. Interrupted time series (ITS) analysis evaluated changes in monthly incidence rates of psychiatric outcomes.

**Results:** During the COVID-19 period a 36% increase was observed in the incidence of depression (95%CI: 25-47), 31% in anxiety (95%CI: 23-39), 20% in stress (95%CI: 13-27), 50% in eating disorders (95%CI: 35-67), 25% in antidepressants (95%CI: 25-33) and 28% in antipsychotics dispensation (95%CI: 18-40). Decreased rate of 26% (95% CI: 0.80-0.88) was observed in ADHD diagnoses and 10% (95% CI: 0.86-0.93) in prescriptions of ADHD agents. The increase was mostly attributed to females in the general Israeli population; nevertheless, a 24% increase in anxiety was seen in males (95%CI: 13-37), 64% in Israeli Arabs (95%CI: 12-140) and 31% in ultra-orthodox (95%CI: 3-67). ITS analysis revealed a significantly higher growth in the incidence of psychiatric outcomes during the COVID-19 period, compared to previous years.

**Conclusions and Relevance:** EHR data of adolescents shows increased incidence rates of mental health diagnoses and medications during the COVID-19 pandemic, specifically identified females as those with the highest mental health burden. Our study highlights that the deteriorating mental health of children should be considered by decision-makers when actions and policies are put in place entering the third year of the pandemic.

**Key Points:** *Question:* Has the COVID-19 pandemic and the strategies to contain it affected adolescents’ mental health?

*Findings:* In this retrospective cohort study of over 200,000 adolescents 12–17 years old, the incidence rates of several measured mental health diagnoses and psychiatric medications increased significantly during the COVID-19 pandemic compared to the period before. This increase was mostly attributed to females.

*Meaning:* This real-world study highlights the deterioration of adolescents’ mental health during the COVID-19 pandemic and suggests that the mental health of this young population should be considered during management and health policy decision making.

## Introduction

The COVID-19 pandemic and measures taken to control its spread have transformed the lives of adolescents, raising concern for their mental health. Although children and adolescents mostly present a milder course of COVID-19 compared to adults ^1,2^ their mental health and wellbeing were negatively impacted during the pandemic ^3^. Current reports have indicated that depression, anxiety and eating disorders have increased significantly since the outbreak of COVID-19 and higher prevalence rates were measured among females ^4–6^, increasing gradually with age ^7^. However, the information regarding the increase in mental health of adolescents during COVID-19 is still limited and is not based on longitudinal follow-up of real-world data population studies. Most of the present studies are based on survey data collected during the early phase of the COVID-19 outbreak. Quantitative study designs, based on real-world data, are needed to assess the changes in mental health of children and adolescents during the COVID-19 pandemic more accurately and compare it to previous years ^7^.

The disruption caused by the pandemic was further exacerbated by the steps that were taken to mitigate it, such as three full lockdowns, social distancing policies and quarantine instructions for those exposed and infected by the SARS-CoV-2 virus. The disruption to the education system affected about 1.8 million students despite significant efforts to deploy distance learning. Previous studies have shown that whenever children are not in their educational routine, they become physically less active, their screen time is much prolonged, they have irregular sleep schedules and their diets are less healthy^8,9^. Furthermore, pandemic stressors such as the threat of the disease, decreased peer interactions, lack of personal space at home, and family’s financial loss may have even more troublesome and enduring impacts on children’s mental health^8^. In this study we quantified the effect of COVID-19 on the incidence of Israeli youth mental health outcomes based on comprehensive EHR data.

## Methods

### STUDY POPULATION AND DESIGN

Data in this study originated from Maccabi Healthcare Services (MHS), the second largest Health Maintenance Organization (HMO) in Israel, which includes 2.5 million insured citizens with longitudinal EHR dated back to 1993. We performed a retrospective cohort study design of adolescents, 12-17 years old (up to their 18^th^ birthday) between November 1^st^, 2016, and October 31^st^, 2021.

### OUTCOMES

We examined incidence rates of several outcomes associated with mental distress. These included five categories of mental health diagnoses: depression (ICD10 F32, F34); anxiety and obsessive-compulsive disorder disorders (ICD10 F41, F42); adjustment and emotional problems, stress-related conditions (ICD10 F43, F93; henceforth denoted as “stress”), eating disorders (ICD10 F50) and attention deficit hyperactivity disorder (ADHD) (ICD10 F90). Furthermore, we assessed four categories of drugs dispensed during those years – antidepressants (ATC code N06A), anxiolytics (ATC code N05B), antipsychotics (ATC code N05A) and psychostimulants, agents used for ADHD and nootropics (ATC code N06B; henceforth denoted as “ADHD agents”). These diagnoses and prescriptions were given by physicians of various specializations (Figure S7).

### DEMOGRAPHIC VARIABLES

We examined trends in mental illness stratified by age sub-groups (12-13, 14-15, 16-17 years old), sex assigned at birth (male, female), sector (general Israeli population, Israeli-Arab, and Ultra-Orthodox Jewish) and socioeconomic status on a scale from 1 to 10 (SES; low 1-3, medium 4-7 and high 8-10). SES and population sectors were determined by the participant’s geo-statistical area of residence using *Points Location Services LTD* (POINTS), which integrates information from the Israeli Central Bureau of Statistics with other socio-economic and demographic data sources^10^. The Israeli Ministry of Health and all four health maintenance organizations use the POINTS data to explore SES and sectors routinely. Individuals with missing sector information (less than 0.01%) and unlisted SES were excluded from the SES sub-analysis (1.1%) but were included in all other analyses. Sex and age were listed for all members.

### STATISTICAL ANALYSIS

Incidence was computed by considering the cohort of all MHS’ members 12-17 years old at the beginning of the year, who did not previously receive a diagnosis, or a medication of the type being considered. The number of members who received the measured diagnosis or medication during the year was counted and normalized by the size of the cohort. Relative Risks (RRs) per 1,000 members and associated 95% confidence intervals (CIs) and p-values were computed to measure the changes and their significance in mental illness trends between each following year and between two time periods: pre-COVID-19 (year 2019 vs 2017) and during COVID-19 (year 2021 vs 2019). Since our data extended up to Oct 31^st^, 2021, each analyzed year started on Nov 1^st^ of the previous year and ended on Oct 31^st^. To present the results of the RRs and 95% CIs we used forest plots.

We used an interrupted time series design (ITS)^11^ to quantify changes in the level and growth in monthly incident rates before and during the COVID-19 pandemic. The interruption was defined on Feb. 27^th^, 2020, the day the first case of COVID-19 was detected in Israel. We used linear regression models and included Fourier terms to model the seasonal factors, with P value <.05 considered statistically significant. Fourier analysis resolves the time dimension variable and allows to identify, quantify, and remove the time-based cycles in the data^12^. Statistical analyses were conducted using Python version 3.7.1 and the statsmodels package version 0.12.

## Ethics declarations

The study protocol was approved by Maccabi Health Services’ institutional review board (MH6-0006-21), and informed consent was waived. The study included retrospective, de-identified data therefore, presented minimal risk and did not adversely affect the rights and welfare of the subjects.

## Results

We evaluated EHR data of 12–17 years old adolescents to assess mental health outcomes before and during the pandemic. For each outcome measured we excluded those with a history of that specific outcome, therefore, the population numbers differed between outcomes. We measured average population sizes of 200,824 in 2017, 207,703 in 2019 and 218,146 in 2021 and this population consisted of 50.4% males on average. The demographic share of the general Israeli population in this cohort was 79.8%, the community of ultra-orthodox Jews was 12.5%, and 7.7% were Israeli Arabs (Table 1). Diagnoses and medication incidence rates for mental health increased over time, however the rise was higher during the COVID-19 period than the pre-COVID-19 period. Overall, the incidence rates of depression increased by 36%, anxiety by 31%, eating disorders by 50% and stress by 20% from 2019 and 2021 (eTable 1).

**Table 1:**
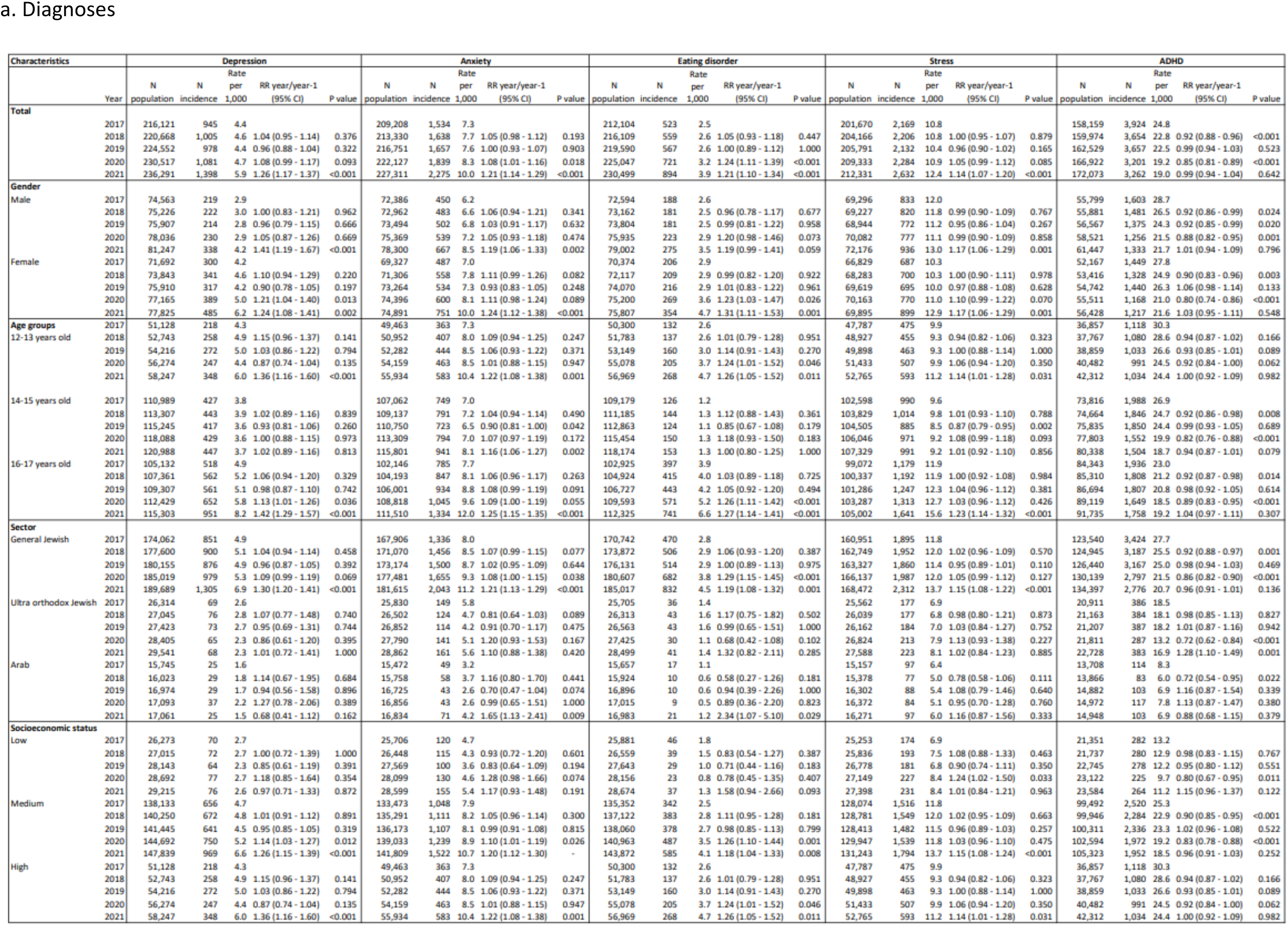
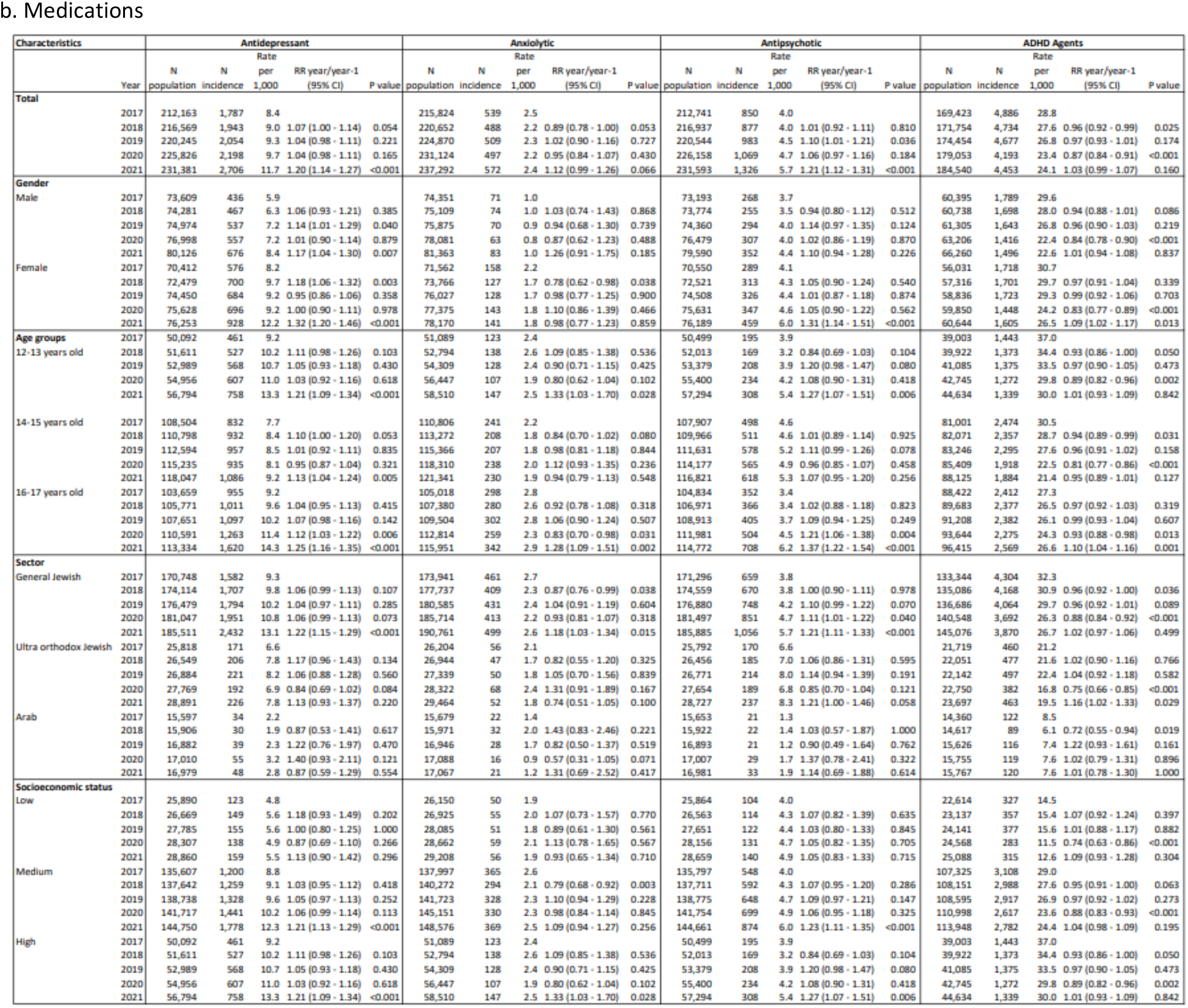
Study population characteristics, incidence rates of mental health diagnoses and medications by year

First, we explored the yearly outlook of all the outcomes tested by plotting the monthly incidence rates of an outcome per 1,000 members for each year from 2017 to 2021. In the beginning of the COVID-19 pandemic (March-April 2020), we observed a drastic decrease in the rates of all diagnoses and medications, corresponding to the first, and strictest, lockdown. However, from May 2020 rates of most diagnoses and medications increased and were high throughout 2021 compared to the years before the pandemic (Figure 1). As the year 2020 included a pre-pandemic period in the beginning of the year, followed by the COVID-19 outbreak in March, a period where access to mental health services was severely disrupted, we analyzed 2021 as the COVID-19 period and 2019 as the pre-pandemic period. To quantify the effect of the COVID-19 period on the mental health of adolescents, we compared the relative risk of incidence in the pre-COVID-19 period (2019 vs 2017) and during the COVID-19 period (2021 vs 2019) (eTable1; Figure 2). The analysis of the COVID-19 period presented a sharp rise in mental health outcomes such as 36% increase in depression (RR=1.36; 95%CI: 1.25-147), 31% in anxiety (RR=1.31; 95% CI: 1.23-1.39), 20% in stress (RR=1.20; 95% CI: 1.13-1.27), 50% in eating disorders (RR=1.50; 95% CI: 1.35-1.67), 25% in antidepressants (RR=1.25; 95% CI: 1.25-1.33) and 28% in antipsychotics (RR=1.28; 95% CI: 1.18-1.40). We found a small, albeit statistically significant, decrease in the rate of ADHD diagnoses (RR=0.84; 95% CI: 0.80-0.88) and corresponding prescriptions of ADHD agents (RR=0.90; 95% CI: 0.86-0.93).

**Figure 1:**
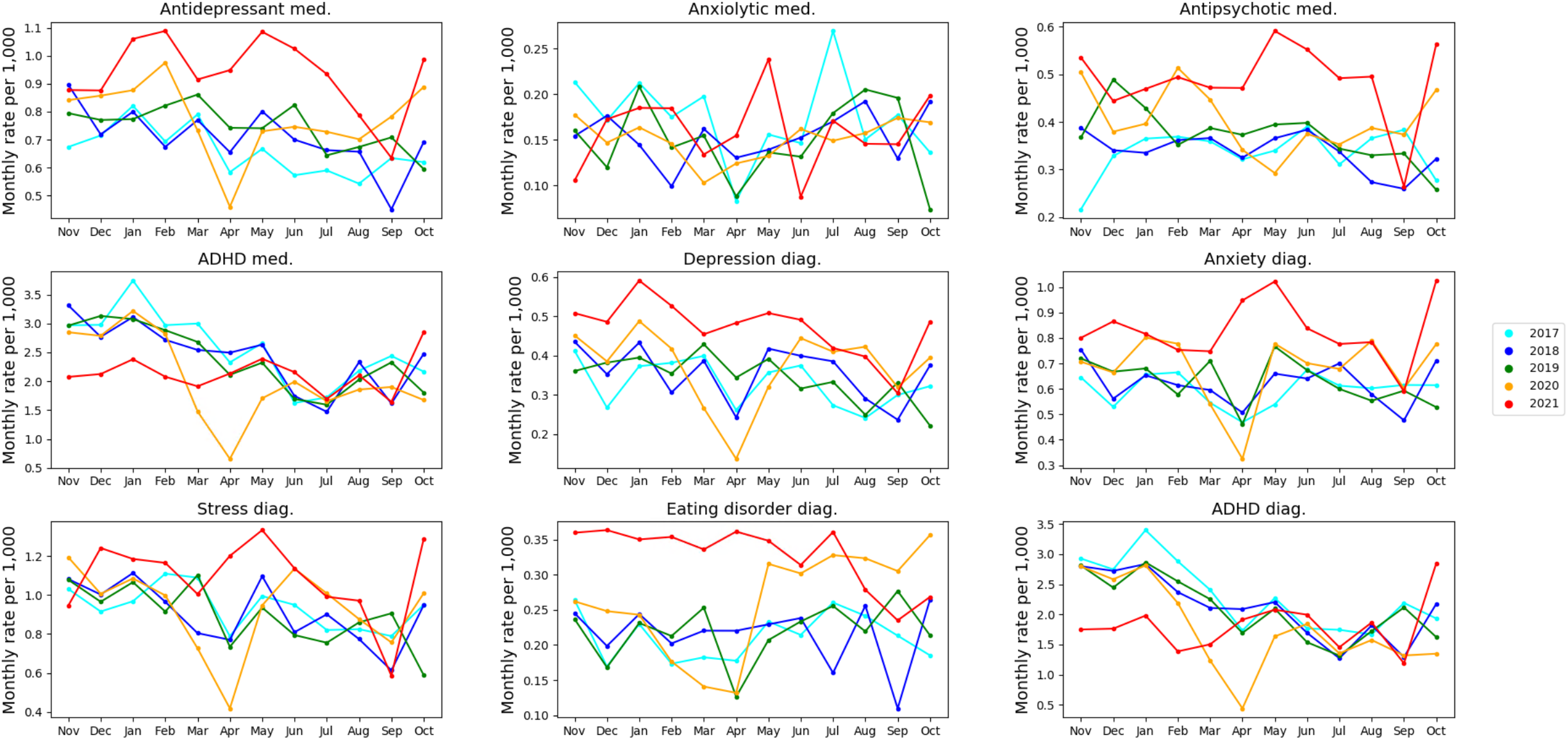
Monthly incidence rates of mental health diagnoses and drug dispensation, comparison by years

**Figure 2:**
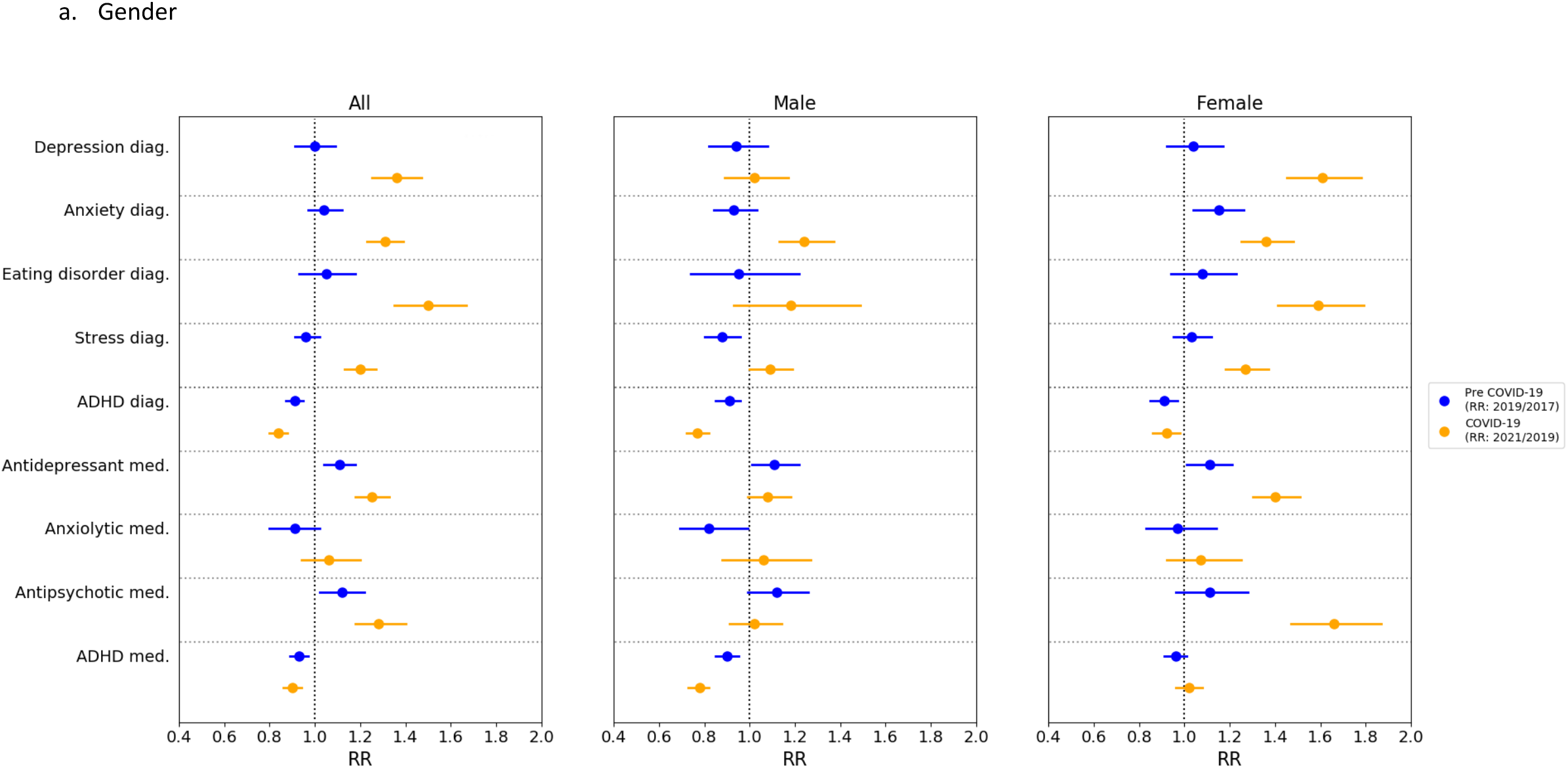
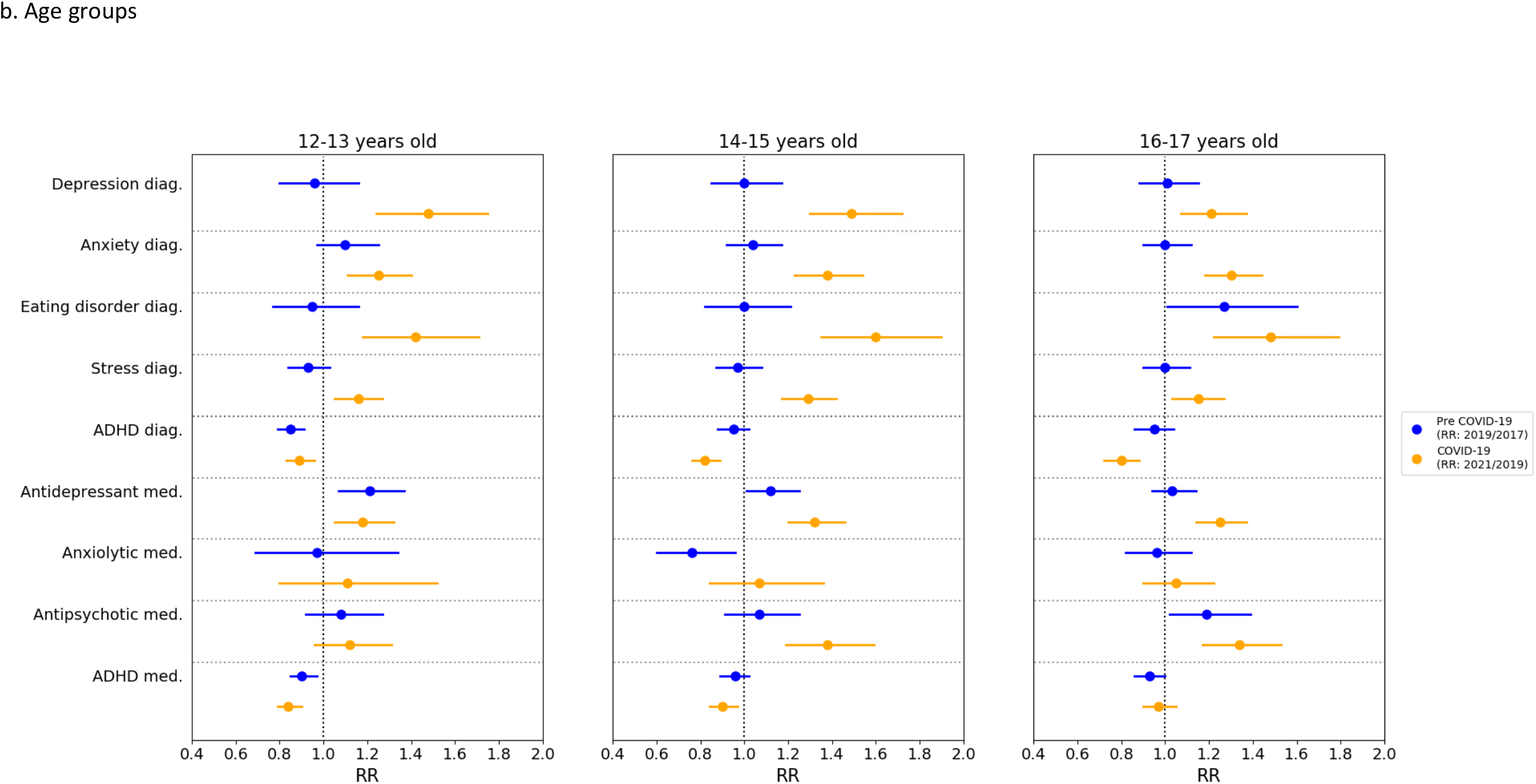
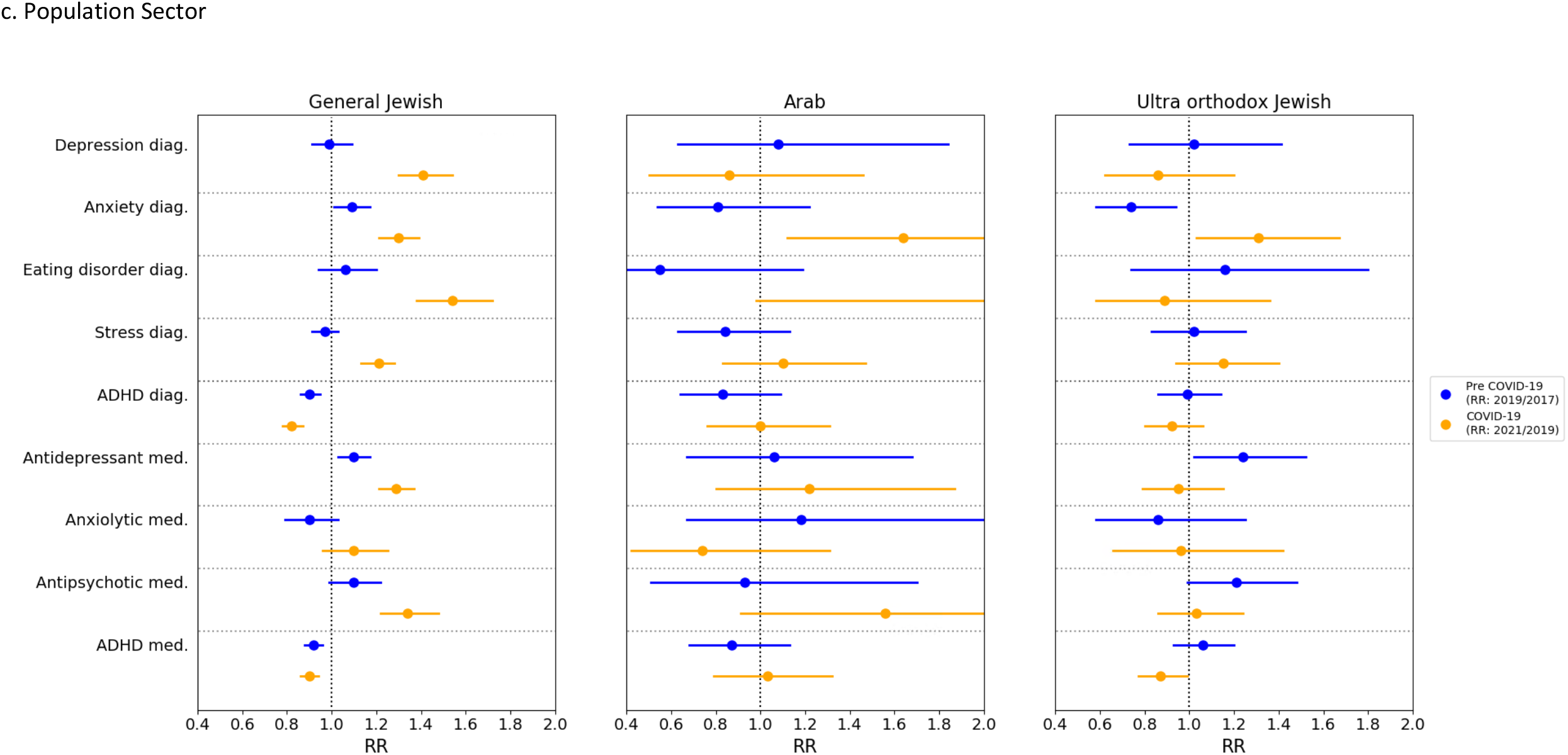
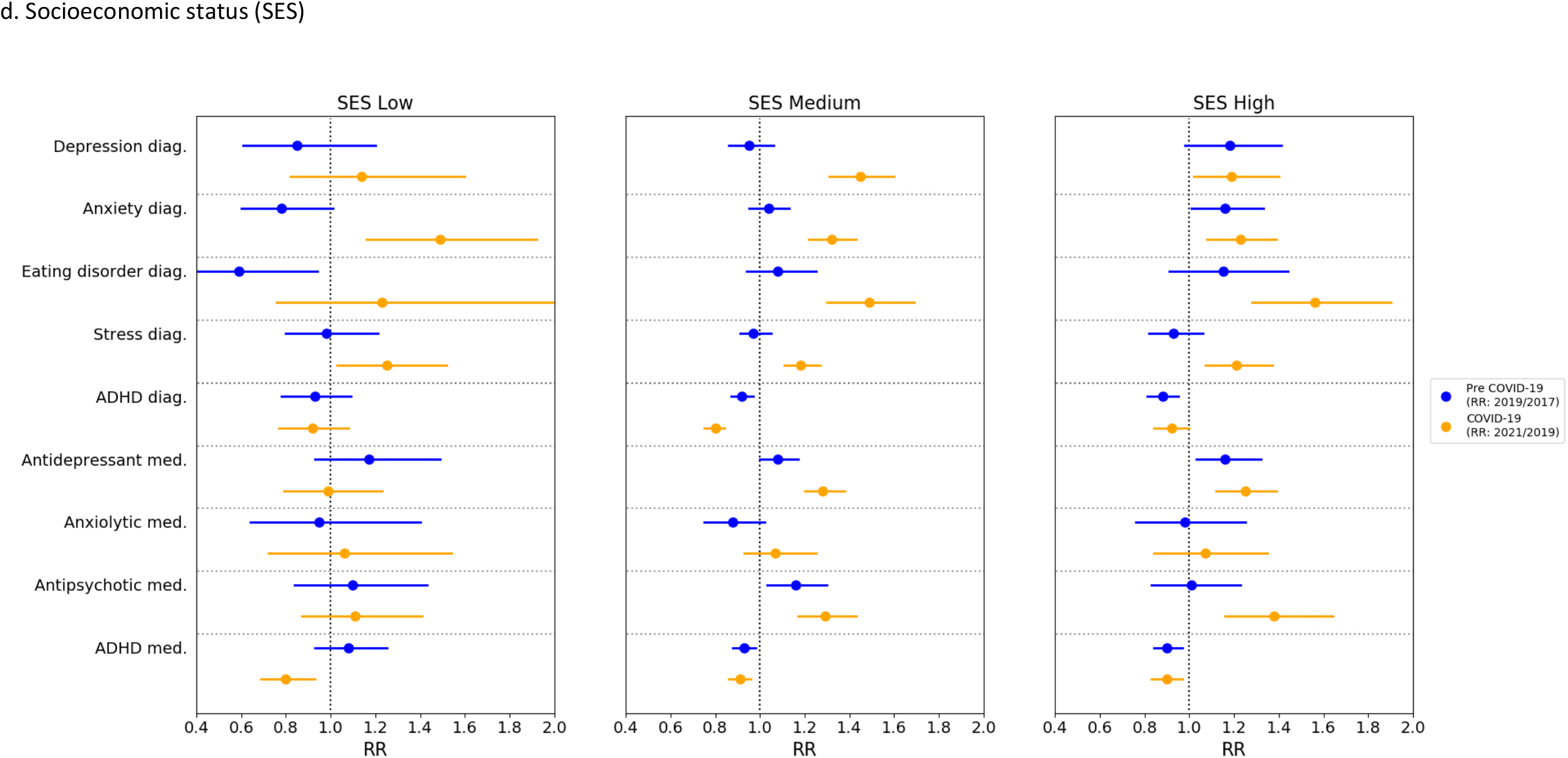
Relative risks (RR) and 95% CI of incidence rates Pre COVID-19 (2019/2017 years; blue) and COVID-19 (2021/2019 years; orange): (a) partition by gender; (b) partition by age; (c) partition by sector; (d) partition by socioeconomic score.

To better describe the specific populations who demonstrated the sharpest increase in incidence of mental health outcomes, we performed additional sub-analyses. In the gender-stratified analyses, most of the increase in incidence rates of psychiatric diagnoses and medications was associated with females, while males generally presented risk rates that were not significantly different from previous years (Figure 2a). While in the pre-COVID-19 period a significant increase among females was only measured in anxiety diagnosis (RR=1.15; 95% CI: 1.04-1.26) and antidepressants dispensation (RR=1.11; 95% CI: 1.01-1.21), during the COVID-19 period we observed significant increases in incidence rates of depression (RR=1.61; 95% CI: 1.45-1.88), anxiety (RR=1.36; 95% CI: 1.25-1.48), stress (RR=1.27; 95% CI: 1.18-1.37), eating disorders (RR=1.59; 95% CI: 1.41-1.79), antidepressants (RR=1.40; 95% CI: 1.3-1.51) and antipsychotics (RR=1.66; 95% CI: 1.47-1.80). The only significant increase measured in males during COVID-19 period was 24% in anxiety (RR=1.24; 95% CI: 1.13-1.37).

Next, we performed a narrower age stratification to evaluate whether a more specific age group was associated with the increase in psychiatric diagnoses or drug dispensation. Age-group analyses have shown a significant increase during COVID-19 period in diagnoses of depression, anxiety, stress and eating disorders among all the groups, with the highest increase observed in the age group of 14-15 years old (Figure 2b). This group presented significant increases in diagnoses of depression (RR=1.49; 95% CI: 1. 1.30 – 1.72), anxiety (RR=1.38; 95% CI: 1.23-1.54), stress (RR=1.29; 95% CI: 1.17-1.42) and eating disorders (RR=1.60; 95% CI: 1.35-1.90). Furthermore, the incidence rates of antidepressants and antipsychotics dispensation had the most pronounced increase among the same age group (RR=1.32; 95% CI: 1.20-1.46 and RR=1.38; 95% CI: 1.19-1.59 respectively).

The Israeli society is composed of different sectors that usually present considerable disparities between them, therefore we sub-analyzed the effect of the COVID-19 period on mental health outcomes of adolescents in the different sectors. The sector-stratified analyses showed that most of the increase in the incidence rates of psychiatric diagnoses and medications dispensation was associated with the general Israeli population. A single significant increase was observed in the Israeli Arab and ultra-orthodox communities in anxiety diagnosis (Figure 2c). The incidence rates of anxiety in the ultra-orthodox community increased by 31% during the pandemic period (RR=1.31; 95% CI: 1.03-1.67) and among Israeli Arabs by 64% (RR=1.64; 95% CI: 1.12-2.40).

The effect of socioeconomic status (SES) on the increased incidence in mental health outcomes in adolescents during the COVID-19 period was also examined. Sub-group analysis by socioeconomic status (Figure 2d) was done by portioning the sample into three groups, low status representing about 12% of the sample, over 60% of the sample with medium status and high status at about 25% of the sample. The medium and high SES groups presented a more distinct change, showing significant incident increases in six outcomes: depression (RR=1.45 95% CI: 1.31-1.60; RR=1.19; 95% CI: 1.02-1.40 respectively), anxiety (RR=1.32 95% CI: 1.22-1.43; RR=1.23; 95% CI: 1.08-1.39), eating disorders (RR=1.49 95% CI: 1.30-1.69; RR=1.56; 95% CI: 1.28-1.90), stress (RR=1.18 95% CI: 1.11-1.27; RR=1.21; 95% CI: 1.07-1.37), antidepressants dispensation (RR=1.28; 95% CI: 1.20-1.38; RR=1.25; 95% CI: 1.12-1.39) and antipsychotics dispensation (RR=1.29; 95% CI: 1.17-1.43; RR=1.38; 95% CI: 1.16-1.64). In the low SES group, the increase was less visible, with only two significantly increased outcomes: anxiety (RR=1.49 95% CI: 1.16-1.92) and stress (RR=1.25 95% CI: 1.03-1.52). The decrease in ADHD agents was observed across all SES groups. Notably, we observed among the high SES group a significant increase in prescription of antidepressants (RR=1.16 95% CI: 1.03-1.32), and among medium SES group a significant increase in prescription of antipsychotics (RR=1.16 95% CI: 1.03-1.30) for the pre-pandemic period.

To enable a more refined analysis examining to what extent the trends during the COVID-19 era are a continuation of past trends and to what extent they break away from them, we performed Interrupted Time Series (ITS) analysis. We evaluated six different models for this analysis which differed in the time periods used to fit the data and the number of interruption points (Figures S1-S6). We observed that following a decline in incidence rates during the first lockdown (from mid-March to the end of April 2020), there was an increase in incidence rates of all diagnoses and medications dispensation, which was significantly higher than the trend in previous years (Figure 3). Varying the analysis by introducing a “gap” period during this first lockdown period, and optionally also during the following month, so as not to be biased by the initial sharp decline, led to qualitatively comparable results (Figures S2-S5). Introducing a second interruption point on March 7^th^, 2021, the day all schools were opened following an extensive vaccination campaign, resulted in a significant decline following that point in the incidence of antidepressant and anxiolytic dispensation as well as of diagnoses of anxiety, stress and eating disorder (eFigure 6 and eTable 2).

**Figure 3:**
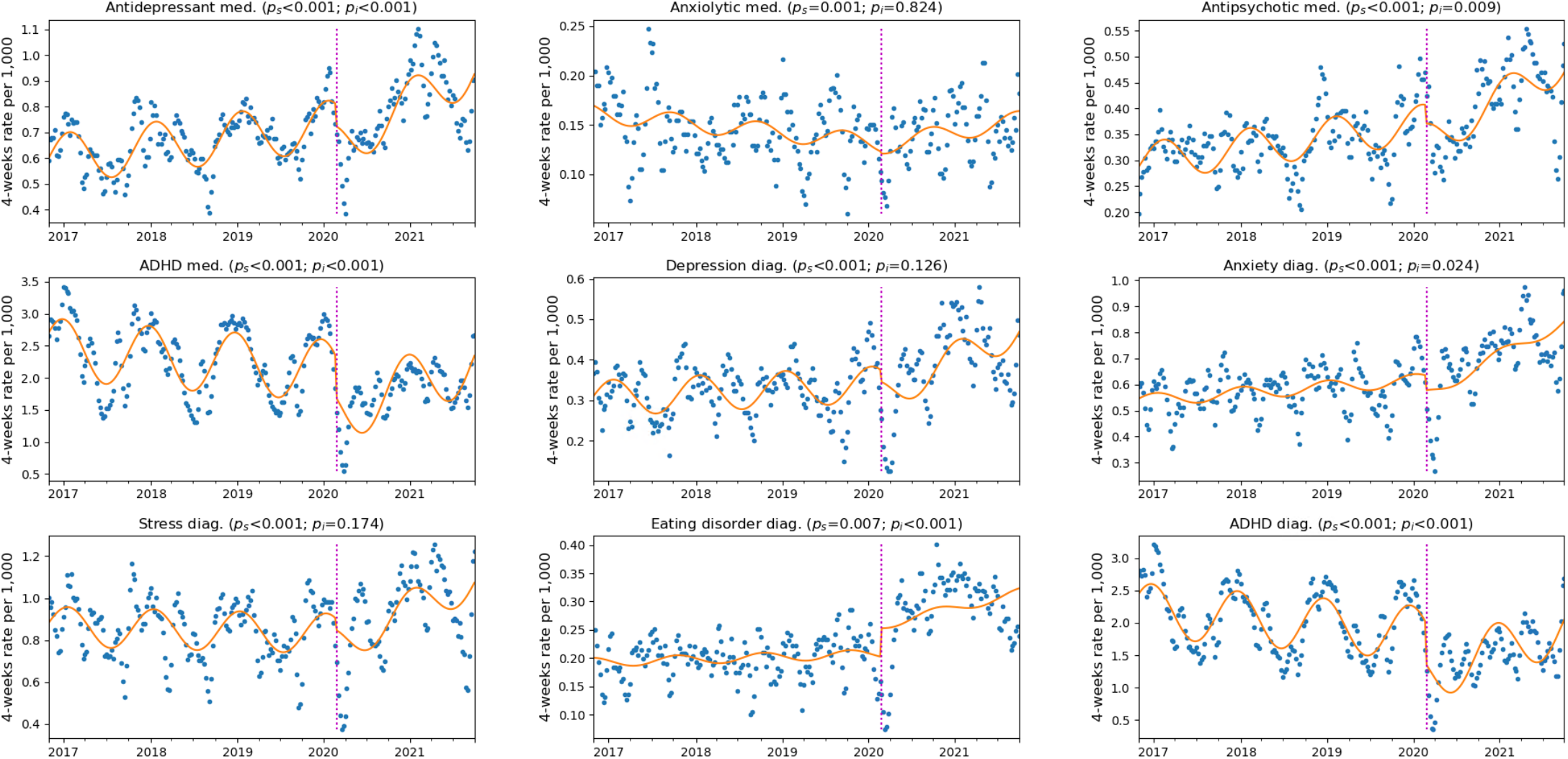
Interrupted Times Series Analysis (ITS) for mental health incidence for 5 years. **Legends:** The magenta line delineates Feb. 27^th^, the date of the first COVID-19 case in Israel. Orange lines depict ITS models with an interruption on this date. Blue dots represent incidence rates for a 4-week period.

## Discussion

The COVID-19 pandemic has taken a toll on the mental health and wellbeing of children and adolescents. While most recent studies used surveys to assess the status of mental health in adolescents ^7,13^, we approached the issue from a quantitative perspective and compared new psychiatric diagnoses and drug dispensation in adolescents before and during the COVID-19 pandemic based on comprehensive EHR data. Consistent with studies showing a sharp increase in reports of mental health problems ^4,7,14^, we observed a significant increase in diagnoses of depression, anxiety, stress and eating disorders during the COVID-19 period compared to previous years. These observations can be attributed to a wide range of stressors that appeared during the pandemic. The increase in depression, anxiety and stress might have been a result of fear of morbidity and mortality (to self or loved ones) from the new unknown illness, excessive media exposure with alarming content, continuous changes in guidelines and restrictions which led to prolonged social isolation, loss of peer interactions and support during school closures (eTable 3), reduced extracurricular and physical activity, disruption of daily routines, decreased hope for the future and loss of pleasure in activities ^15–17^. The introduction of new distance learning technologies imposed new challenges involving constant self-observation through cameras and different academic success evaluation, compromising adolescents’ self-esteem. When Eating disorders are associated with body dissatisfaction, poor self-esteem and depression^18^, an illusion of control over the body may have been more readily triggered by the uncertainty of the new reality. While the increase in psychiatric drug dispensation was mostly aligned with the corresponding diagnoses, the increase in dispensation of antipsychotic drugs was not associated with any specific diagnosis measured in this study and was particularly pronounced in females, might indicate incidences of self-harm and personality disorders^19^. Among the reasons for increased incidence of psychiatric outcomes, we should also consider the extended time periods adolescents spent at home with their immediate family that may have promoted enhanced parental awareness and led to increased legitimacy to discuss mental distress during these times ^20^.

While rates of depression and anxiety increased during the COVID-19 period in this study, we observed a reduction in the rates of ADHD diagnoses and prescription medications. As reports in recent years have shown an overall rise in ADHD diagnoses and medications ^21,22^, one explanation of our findings might be that ADHD diagnoses and medication use are seasonal, with adherence levels reported to be higher during the school year than during school vacations^23–26^. Since the COVID-19 period was characterized by intermittent closure of schools, this observed reduction in diagnoses and drug use is in line with the seasonal decrease during a normal school year’s summer break. Furthermore, lower attendance in classes reduced student numbers which may have created a better environment for those with concentration difficulties. This reality might have also increased teachers’ tolerance to class absence, lack of task completion and behavioral problems, therefore less inclined to refer adolescents to ADHD evaluation. These findings may suggest under-diagnosed and untreated ADHD in adolescents putting them at risk for more serious outcomes, such as increased rates of criminal activity, accidents as well as anxiety and depression^27^.

The most significant finding in this study is the greater risk of adolescent females diagnosed with a variety of mental disorders for the first time during the pandemic compared to the risk observed for males. In this EHR analysis, risk for anxiety was the only significant mental outcome that was increased among males. These findings are consistent with previous studies which suggested that loneliness was associated with elevated depression symptoms in females and with elevated social anxiety in males ^28,29^. Other studies found both increased depressive and anxiety symptoms among females^4–7^. The World Health Organization (WHO) reported lower levels of mental health and life satisfaction among females compared to males^30^. Gender differences in mental disorders are known and consistent in the field of psychiatry, but the reasons for these differences are still not clear enough^31^. The potential risk factors could be the influence of sex hormones, higher rates of interpersonal stressors, females’ lower baseline self-esteem and higher tendency for body shame, exposure to stress associated with lack of gender equality and discrimination and higher chance of experiencing interpersonal violence^31–33^. While this could imply that females are more sensitive to the effects of the pandemic, this can also be attributed to higher tendency of females sharing their mental distress with their parents or physicians and getting the diagnosis and treatment for their condition. The apparent lack of increase among males might be because they are more frequently referred to mental health services by their school following behavioral issues. As schools were closed intermittently during the analyzed period, potential increase due to the pandemic may have been offset by the decrease in school referrals^29^. Importantly, mental distress among male adolescents may manifest in ways which are not directly reported in the EHR, such as violence, dropout, and substance abuse.

Stratifying by age, our findings show that there was an increase in mental illness in all age groups, more pronounced among 14-15-year-olds. Yet, as described in previous studies^7^, we found that the incidence rates themselves, of depression, anxiety, and the associated medication, are highest among 16–17-year-olds. High rates among older adolescents may be due to changes associated with puberty, a response to the stress of lockdowns and the global pandemic, and lack of socialization with peers which is particularly important at this age. In younger children increased anxiety and depression may be more readily attributed to changes in routine^34,35^. Comparing the sectors in the Israeli population showed that different circumstances and lifestyle during the COVID-19 period are associated with different mental health outcomes in adolescents. In the ultra-orthodox community, we observed a significant increase only in the diagnosis of anxiety (31%). Ultra-orthodox Jews, accounting for 12% of the Israeli population, form of a closely-knit religious communities, living by strict Jewish laws and tradition^36^. In this sector, the pandemic caused tension between governmental instructions and instructions from prominent community leaders who advocated keeping schools open for holy studies ^37,38^. Many media venues such as TV, internet, and secular newspapers are not used in this sector, potentially buffering them from the exacerbating impact of reports and discussions in these venues. Moreover, while in recent years there is a growing openness and legitimacy for the discussion of mental health in this community, utilization of mental health services is still much lower than in the general Israeli population^37,39^. Similar to the Ultra-Orthodox sector, in the Israeli-Arab sector we observed a significant increase only in the diagnosis of anxiety (64%). This sector, which accounts for 20% of the total Israeli population, utilizes mental health services at a much lower rate than the rest of the population, possibly due to lower access to healthcare services as many reside in peripheral areas, as well as negative cultural perception and stigma associated with mental health problems^40,41^.

A significant increase in mental health diagnoses during the COVID-19 pandemic period was observed across different SES. Differently from previous studies reporting that children and adolescents who grow up in families with lower SES have more symptoms of anxiety and depression^42–44^, the incidence rates of most diagnoses in this study increased with higher SES. Distinctly in Israel, the Israeli-Arab and ultra-orthodox communities are associated with lower SES^37^ and as mentioned earlier, in these sectors the rates of diagnoses related to mental status are lower than the rest of the population for cultural, religious, and other reasons. Although lower SES is often associated with poor mental health, other mediators might affect this association. Social engagement with friends and family is linked to better mental health and affects the link between SES and mental health^45^. Ultra-orthodox communities in Israel are usually of lower SES, nevertheless they are socially active environments which provide members with social and spiritual support^46^. The community settings offer psychosocial engagement and continuous assistance which results in better health and mental health that would be expected based on their SES^46,47^.

Our study has several limitations. While the findings in this study are clear and consistent with other studies, the reported incidence rates are probably an underestimation of the actual numbers. Many adolescents are diagnosed and treated by mental health professionals in private clinics outside their HMO, and such diagnoses are not recorded in their EHR. Psychiatric drugs prescribed outside the HMO are also not recorded and were not included in this analysis. Furthermore, there is a long standby time for mental health services, starting at an average of three months before an initial assessment, and several additional months before receiving treatment^48^ - so not all those who seek and need help are included in this study. Our analysis addressed these limitations by comparing the risk ratios of the outcomes, measuring the difference in new mental health diagnoses and dispensation within the HMO in different time periods.

In this study, the rates of new cases of mental health diagnoses and drug dispensation were determined in individuals that had no history of the specific measured outcome. However, mental health often presents multiple comorbidities at different times in patients’ history, i.e. depression may appear before anxiety diagnosis or anti-depressants dispensation. Future studies could explore how pre-COVID mental health diagnoses and prescriptions affected new mental health outcomes during the pandemic.

## Conclusions

To the best of our knowledge, this study is the first data-driven quantitative estimation of the mental health burden on adolescents during the COVID-19 pandemic. This observational cohort study showed a significant increase in mental health diagnoses and in psychiatric drugs dispensation during the COVID-19 period compared to the corresponding pre-COVID period. Our findings highlight the specific sub-populations which need to be considered when deciding on policies and promotion of adolescent resilience. These findings should warrant similar studies in other geographical areas and in other age groups. Strategies to address the deteriorating mental health of adolescents during the COVID-19 pandemic and prevent further deterioration as the disease continues to spread should be a priority.

## Data Availability

The data that support the findings of this study originate from Maccabi Health Services. Restrictions apply to the availability of these data and they are therefore not publicly available. Due to restrictions, these data can be accessed only by request to the authors and/or Maccabi Health Services.

## Acknowledgments

We thank Yossi Levi, Yair Goldberg and Stephen Levine for helpful comments and discussion on the Interrupted Time Series methodology. Chen Yanover and Tal El-Hai are acknowledged for their insightful methodological suggestions and illuminating discussions.

