## Supplementary material for "Data-driven assessment of adolescents’ mental health during the COVID-19 pandemic"

**eTable 1** - Study population characteristics, incidence rates of mental health diagnoses and medications by year (3 years)

a. Diagnoses

| Characteristics | | Depression | | | Anxiety | | | Eating disorder | | | Stress | | | ADHD | | |
|---|---|---|---|---|---|---|---|---|---|---|---|---|---|---|---|---|
| | Year | Rate per 1,000 | RR year/year-2 (95%CI) | P value | Rate per 1,000 | RR year/year-2 (95%CI) | P value | Rate per 1,000 | RR year/year-2 (95%CI) | P value | Rate per 1,000 | RR year/year-2 (95%CI) | P value | Rate per 1,000 | RR year/year-2 (95%CI) | P value |
| **Total** | | | | | | | | | | | | | | | | |
| | 2017 | 4.4 | | | 7.3 | | | 2.5 | | | 10.8 | | | 24.8 | | |
| | 2019 | 4.4 | 1.00 (0.91 - 1.09) | 0.945 | 7.6 | 1.04 (0.97 - 1.12) | 0.241 | 2.6 | 1.05 (0.93 - 1.18) | 0.448 | 10.4 | 0.96 (0.91 - 1.02) | 0.220 | 22.5 | 0.91 (0.87 - 0.95) | <0.001 |
| | 2021 | 5.9 | 1.36 (1.25 - 1.47) | <0.001 | 10.0 | 1.31 (1.23 - 1.39) | <0.001 | 3.9 | 1.50 (1.35 - 1.67) | <0.001 | 12.4 | 1.20 (1.13 - 1.27) | <0.001 | 19.0 | 0.84 (0.80 - 0.88) | <0.001 |
| **Age groups** | | | | | | | | | | | | | | | | |
| 12-13 years old | 2017 | 2.9 | | | 6.2 | | | 2.6 | | | 12.0 | | | 28.7 | | |
| | 2019 | 2.8 | 0.96 (0.80 - 1.16) | 0.700 | 6.8 | 1.10 (0.97 - 1.25) | 0.152 | 2.5 | 0.95 (0.77 - 1.16) | 0.603 | 11.2 | 0.93 (0.84 - 1.03) | 0.160 | 24.3 | 0.85 (0.79 - 0.91) | <0.001 |
| | 2021 | 4.2 | 1.48 (1.24 - 1.75) | <0.001 | 8.5 | 1.25 (1.11 - 1.40) | <0.001 | 3.5 | 1.42 (1.18 - 1.71) | <0.001 | 13.0 | 1.16 (1.05 - 1.27) | 0.003 | 21.7 | 0.89 (0.83 - 0.96) | 0.003 |
| 14-15 years old | 2017 | 4.2 | | | 7.0 | | | 2.9 | | | 10.3 | | | 27.8 | | |
| | 2019 | 4.2 | 1.00 (0.85 - 1.17) | 1.000 | 7.3 | 1.04 (0.92 - 1.17) | 0.572 | 2.9 | 1.00 (0.82 - 1.21) | 1.000 | 10.0 | 0.97 (0.87 - 1.08) | 0.589 | 26.3 | 0.95 (0.88 - 1.02) | 0.141 |
| | 2021 | 6.2 | 1.49 (1.30 - 1.72) | <0.001 | 10.0 | 1.38 (1.23 - 1.54) | <0.001 | 4.7 | 1.60 (1.35 - 1.90) | <0.001 | 12.9 | 1.29 (1.17 - 1.42) | <0.001 | 21.6 | 0.82 (0.76 - 0.89) | <0.001 |
| 16-17 years old | 2017 | 6.1 | | | 8.8 | | | 1.9 | | | 9.9 | | | 17.4 | | |
| | 2019 | 6.1 | 1.01 (0.88 - 1.15) | 0.919 | 8.9 | 1.00 (0.90 - 1.12) | 0.977 | 2.4 | 1.27 (1.01 - 1.60) | 0.043 | 9.9 | 1.00 (0.90 - 1.11) | 1.000 | 16.4 | 0.95 (0.86 - 1.04) | 0.252 |
| | 2021 | 7.4 | 1.21 (1.07 - 1.37) | 0.002 | 11.6 | 1.30 (1.18 - 1.44) | <0.001 | 3.5 | 1.48 (1.22 - 1.79) | <0.001 | 11.3 | 1.15 (1.03 - 1.27) | 0.009 | 13.1 | 0.80 (0.72 - 0.88) | <0.001 |
| **Gender** | | | | | | | | | | | | | | | | |
| Male | 2017 | 3.8 | | | 7.0 | | | 1.2 | | | 9.6 | | | 26.9 | | |
| | 2019 | 3.6 | 0.94 (0.82 - 1.08) | 0.389 | 6.5 | 0.93 (0.84 - 1.03) | 0.191 | 1.1 | 0.95 (0.74 - 1.22) | 0.705 | 8.5 | 0.88 (0.80 - 0.96) | 0.005 | 24.4 | 0.91 (0.85 - 0.96) | 0.002 |
| | 2021 | 3.7 | 1.02 (0.89 - 1.17) | 0.785 | 8.1 | 1.24 (1.13 - 1.37) | <0.001 | 1.3 | 1.18 (0.93 - 1.49) | 0.186 | 9.2 | 1.09 (1.00 - 1.19) | 0.064 | 18.7 | 0.77 (0.72 - 0.82) | <0.001 |
| Female | 2017 | 4.9 | | | 7.7 | | | 3.9 | | | 11.9 | | | 23.0 | | |
| | 2019 | 5.1 | 1.04 (0.92 - 1.17) | 0.522 | 8.8 | 1.15 (1.04 - 1.26) | 0.005 | 4.2 | 1.08 (0.94 - 1.23) | 0.300 | 12.3 | 1.03 (0.95 - 1.12) | 0.402 | 20.8 | 0.91 (0.85 - 0.97) | 0.003 |
| | 2021 | 8.2 | 1.61 (1.45 - 1.78) | <0.001 | 12.0 | 1.36 (1.25 - 1.48) | <0.001 | 6.6 | 1.59 (1.41 - 1.79) | <0.001 | 15.6 | 1.27 (1.18 - 1.37) | <0.001 | 19.2 | 0.92 (0.86 - 0.98) | 0.012 |
| **Sector** | | | | | | | | | | | | | | | | |
| General Jewish | 2017 | 4.9 | | | 8.0 | | | 2.8 | | | 11.8 | | | 27.7 | | |
| | 2019 | 4.9 | 0.99 (0.91 - 1.09) | 0.923 | 8.7 | 1.09 (1.01 - 1.17) | 0.024 | 2.9 | 1.06 (0.94 - 1.20) | 0.371 | 11.4 | 0.97 (0.91 - 1.03) | 0.309 | 25.0 | 0.90 (0.86 - 0.95) | <0.001 |
| | 2021 | 6.9 | 1.41 (1.30 - 1.54) | <0.001 | 11.2 | 1.30 (1.21 - 1.39) | <0.001 | 4.5 | 1.54 (1.38 - 1.72) | <0.001 | 13.7 | 1.21 (1.13 - 1.28) | <0.001 | 20.7 | 0.82 (0.78 - 0.87) | <0.001 |
| Ultra orthodox Jewish | 2017 | 2.6 | | | 5.8 | | | 1.4 | | | 6.9 | | | 18.5 | | |
| | 2019 | 2.7 | 1.02 (0.73 - 1.41) | 0.933 | 4.2 | 0.74 (0.58 - 0.94) | 0.013 | 1.6 | 1.16 (0.74 - 1.80) | 0.574 | 7.0 | 1.02 (0.83 - 1.25) | 0.916 | 18.2 | 0.99 (0.86 - 1.14) | 0.885 |
| | 2021 | 2.3 | 0.86 (0.62 - 1.20) | 0.400 | 5.6 | 1.31 (1.03 - 1.67) | 0.025 | 1.4 | 0.89 (0.58 - 1.36) | 0.662 | 8.1 | 1.15 (0.94 - 1.40) | 0.164 | 16.9 | 0.92 (0.80 - 1.06) | 0.275 |
| Arab | 2017 | 1.6 | | | 3.2 | | | 1.1 | | | 6.4 | | | 8.3 | | |
| | 2019 | 1.7 | 1.08 (0.63 - 1.84) | 0.892 | 2.6 | 0.81 (0.54 - 1.22) | 0.347 | 0.6 | 0.55 (0.25 - 1.19) | 0.128 | 5.4 | 0.84 (0.63 - 1.13) | 0.268 | 6.9 | 0.83 (0.64 - 1.09) | 0.195 |
| | 2021 | 1.5 | 0.86 (0.50 - 1.46) | 0.589 | 4.2 | 1.64 (1.12 - 2.40) | 0.011 | 1.2 | 2.09 (0.98 - 4.44) | 0.071 | 6.0 | 1.10 (0.83 - 1.47) | 0.508 | 6.9 | 1.00 (0.76 - 1.31) | 1.000 |
| **Socioeconomic status** | | | | | | | | | | | | | | | | |
| Low | 2017 | 2.7 | | | 4.7 | | | 1.8 | | | 6.9 | | | 13.2 | | |
| | 2019 | 2.3 | 0.85 (0.61 - 1.20) | 0.387 | 3.6 | 0.78 (0.60 - 1.01) | 0.068 | 1.0 | 0.59 (0.37 - 0.94) | 0.028 | 6.8 | 0.98 (0.80 - 1.21) | 0.873 | 12.2 | 0.93 (0.78 - 1.09) | 0.372 |
| | 2021 | 2.6 | 1.14 (0.82 - 1.60) | 0.447 | 5.4 | 1.49 (1.16 - 1.92) | 0.002 | 1.3 | 1.23 (0.76 - 2.00) | 0.460 | 8.4 | 1.25 (1.03 - 1.52) | 0.026 | 11.2 | 0.92 (0.77 - 1.08) | 0.320 |
| Medium | 2017 | 4.7 | | | 7.9 | | | 2.5 | | | 11.8 | | | 25.3 | | |
| | 2019 | 4.5 | 0.95 (0.86 - 1.06) | 0.404 | 8.1 | 1.04 (0.95 - 1.13) | 0.424 | 2.7 | 1.08 (0.94 - 1.25) | 0.296 | 11.5 | 0.97 (0.91 - 1.05) | 0.497 | 23.3 | 0.92 (0.87 - 0.97) | 0.003 |
| | 2021 | 6.6 | 1.45 (1.31 - 1.60) | <0.001 | 10.7 | 1.32 (1.22 - 1.43) | <0.001 | 4.1 | 1.49 (1.30 - 1.69) | <0.001 | 13.7 | 1.18 (1.11 - 1.27) | <0.001 | 18.5 | 0.80 (0.75 - 0.84) | <0.001 |
| High | 2017 | 4.3 | | | 7.3 | | | 2.6 | | | 9.9 | | | 30.3 | | |
| | 2019 | 5.0 | 1.18 (0.98 - 1.41) | 0.077 | 8.5 | 1.16 (1.01 - 1.33) | 0.040 | 3.0 | 1.15 (0.91 - 1.44) | 0.265 | 9.3 | 0.93 (0.82 - 1.06) | 0.294 | 26.6 | 0.88 (0.81 - 0.95) | 0.002 |
| | 2021 | 6.0 | 1.19 (1.02 - 1.40) | 0.033 | 10.4 | 1.23 (1.08 - 1.39) | 0.001 | 4.7 | 1.56 (1.28 - 1.90) | <0.001 | 11.2 | 1.21 (1.07 - 1.37) | 0.002 | 24.4 | 0.92 (0.84 - 1.00) | 0.055 |

b. Medications

| Characteristics | Year | Antidepressant | | | Anxiolytic | | | Antipsychotic | | | ADHD Agents | | |
|---|---|---|---|---|---|---|---|---|---|---|---|---|---|
| | | per 1,000 | RR year/year-2 (95%CI) | P value | per 1,000 | RR year/year-2 (95%CI) | P value | per 1,000 | RR year/year-2 (95%CI) | P value | per 1,000 | RR year/year-2 (95%CI) | P value |
| **Total** | | | | | | | | | | | | | |
| | 2017 | 8.4 | | | 2.5 | | | 4.0 | | | 28.8 | | |
| | 2019 | 9.3 | 1.11 (1.04 - 1.18) | 0.002 | 2.3 | 0.91 (0.80 - 1.02) | 0.115 | 4.5 | 1.12 (1.02 - 1.22) | 0.019 | 26.8 | 0.93 (0.89 - 0.97) | <0.001 |
| | 2021 | 11.7 | 1.25 (1.18 - 1.33) | <0.001 | 2.4 | 1.06 (0.94 - 1.20) | 0.315 | 5.7 | 1.28 (1.18 - 1.40) | <0.001 | 24.1 | 0.90 (0.86 - 0.94) | <0.001 |
| **Age groups** | | | | | | | | | | | | | |
| 12-13 years old | 2017 | 5.9 | | | 1.0 | | | 3.7 | | | 29.6 | | |
| | 2019 | 7.2 | 1.21 (1.07 - 1.37) | 0.003 | 0.9 | 0.97 (0.69 - 1.34) | 0.866 | 4.0 | 1.08 (0.92 - 1.27) | 0.375 | 26.8 | 0.90 (0.85 - 0.97) | 0.003 |
| | 2021 | 8.4 | 1.18 (1.05 - 1.32) | 0.005 | 1.0 | 1.11 (0.80 - 1.52) | 0.571 | 4.4 | 1.12 (0.96 - 1.31) | 0.156 | 22.6 | 0.84 (0.79 - 0.90) | <0.001 |
| 14-15 years old | 2017 | 8.2 | | | 2.2 | | | 4.1 | | | 30.7 | | |
| | 2019 | 9.2 | 1.12 (1.01 - 1.25) | 0.042 | 1.7 | 0.76 (0.60 - 0.96) | 0.024 | 4.4 | 1.07 (0.91 - 1.25) | 0.419 | 29.3 | 0.96 (0.89 - 1.02) | 0.171 |
| | 2021 | 12.2 | 1.32 (1.20 - 1.46) | <0.001 | 1.8 | 1.07 (0.84 - 1.36) | 0.583 | 6.0 | 1.38 (1.19 - 1.59) | <0.001 | 26.5 | 0.90 (0.84 - 0.97) | 0.003 |
| 16-17 years old | 2017 | 11.4 | | | 4.4 | | | 4.2 | | | 26.0 | | |
| | 2019 | 11.8 | 1.03 (0.94 - 1.14) | 0.514 | 4.3 | 0.96 (0.82 - 1.12) | 0.629 | 5.1 | 1.19 (1.02 - 1.39) | 0.026 | 24.1 | 0.93 (0.86 - 1.00) | 0.051 |
| | 2021 | 14.7 | 1.25 (1.14 - 1.37) | <0.001 | 4.5 | 1.05 (0.90 - 1.22) | 0.532 | 6.8 | 1.34 (1.17 - 1.53) | <0.001 | 23.5 | 0.97 (0.90 - 1.05) | 0.456 |
| **Gender** | | | | | | | | | | | | | |
| Male | 2017 | 7.7 | | | 2.2 | | | 4.6 | | | 30.5 | | |
| | 2019 | 8.5 | 1.11 (1.01 - 1.22) | 0.031 | 1.8 | 0.82 (0.69 - 0.99) | 0.042 | 5.2 | 1.12 (0.99 - 1.26) | 0.062 | 27.6 | 0.90 (0.85 - 0.95) | <0.001 |
| | 2021 | 9.2 | 1.08 (0.99 - 1.18) | 0.075 | 1.9 | 1.06 (0.88 - 1.27) | 0.598 | 5.3 | 1.02 (0.91 - 1.14) | 0.728 | 21.4 | 0.78 (0.73 - 0.82) | <0.001 |
| Female | 2017 | 9.2 | | | 2.8 | | | 3.4 | | | 27.3 | | |
| | 2019 | 10.2 | 1.11 (1.01 - 1.21) | 0.022 | 2.8 | 0.97 (0.83 - 1.14) | 0.744 | 3.7 | 1.11 (0.96 - 1.28) | 0.166 | 26.1 | 0.96 (0.91 - 1.01) | 0.128 |
| | 2021 | 14.3 | 1.40 (1.30 - 1.51) | <0.001 | 2.9 | 1.07 (0.92 - 1.25) | 0.407 | 6.2 | 1.66 (1.47 - 1.87) | <0.001 | 26.6 | 1.02 (0.96 - 1.08) | 0.480 |
| **Sector** | | | | | | | | | | | | | |
| General Jewish | 2017 | 9.3 | | | 2.7 | | | 3.8 | | | 32.3 | | |
| | 2019 | 10.2 | 1.10 (1.03 - 1.17) | 0.007 | 2.4 | 0.90 (0.79 - 1.03) | 0.123 | 4.2 | 1.10 (0.99 - 1.22) | 0.078 | 29.7 | 0.92 (0.88 - 0.96) | <0.001 |
| | 2021 | 13.1 | 1.29 (1.21 - 1.37) | <0.001 | 2.6 | 1.10 (0.96 - 1.25) | 0.168 | 5.7 | 1.34 (1.22 - 1.48) | <0.001 | 26.7 | 0.90 (0.86 - 0.94) | <0.001 |
| Ultra orthodox Jewish | 2017 | 6.6 | | | 2.1 | | | 6.6 | | | 21.2 | | |
| | 2019 | 8.2 | 1.24 (1.02 - 1.52) | 0.033 | 1.8 | 0.86 (0.58 - 1.25) | 0.438 | 8.0 | 1.21 (0.99 - 1.48) | 0.065 | 22.4 | 1.06 (0.93 - 1.20) | 0.378 |
| | 2021 | 7.8 | 0.95 (0.79 - 1.15) | 0.602 | 1.8 | 0.96 (0.66 - 1.42) | 0.921 | 8.3 | 1.03 (0.86 - 1.24) | 0.741 | 19.5 | 0.87 (0.77 - 0.99) | 0.031 |
| Arab | 2017 | 2.2 | | | 1.4 | | | 1.3 | | | 8.5 | | |
| | 2019 | 2.3 | 1.06 (0.67 - 1.68) | 0.816 | 1.7 | 1.18 (0.67 - 2.06) | 0.575 | 1.2 | 0.93 (0.51 - 1.70) | 0.878 | 7.4 | 0.87 (0.68 - 1.13) | 0.298 |
| | 2021 | 2.8 | 1.22 (0.80 - 1.87) | 0.391 | 1.2 | 0.74 (0.42 - 1.31) | 0.32 | 1.9 | 1.56 (0.91 - 2.70) | 0.133 | 7.6 | 1.03 (0.79 - 1.32) | 0.896 |
| **Socioeconomic status** | | | | | | | | | | | | | |
| Low | 2017 | 4.8 | | | 1.9 | | | 4.0 | | | 14.5 | | |
| | 2019 | 5.6 | 1.17 (0.93 - 1.49) | 0.186 | 1.8 | 0.95 (0.64 - 1.40) | 0.842 | 4.4 | 1.10 (0.84 - 1.43) | 0.505 | 15.6 | 1.08 (0.93 - 1.25) | 0.323 |
| | 2021 | 5.5 | 0.99 (0.79 - 1.23) | 0.955 | 1.9 | 1.06 (0.72 - 1.54) | 0.847 | 4.9 | 1.11 (0.87 - 1.41) | 0.421 | 12.6 | 0.80 (0.69 - 0.93) | 0.004 |
| Medium | 2017 | 8.8 | | | 2.6 | | | 4.0 | | | 29.0 | | |
| | 2019 | 9.6 | 1.08 (1.00 - 1.17) | 0.048 | 2.3 | 0.88 (0.75 - 1.02) | 0.08 | 4.7 | 1.16 (1.03 - 1.30) | 0.012 | 26.9 | 0.93 (0.88 - 0.98) | 0.003 |
| | 2021 | 12.3 | 1.28 (1.20 - 1.38) | <0.001 | 2.5 | 1.07 (0.93 - 1.25) | 0.363 | 6.0 | 1.29 (1.17 - 1.43) | <0.001 | 24.4 | 0.91 (0.86 - 0.96) | <0.001 |
| High | 2017 | 9.2 | | | 2.4 | | | 3.9 | | | 37.0 | | |
| | 2019 | 10.7 | 1.16 (1.03 - 1.32) | 0.014 | 2.4 | 0.98 (0.76 - 1.25) | 0.899 | 3.9 | 1.01 (0.83 - 1.23) | 0.960 | 33.5 | 0.90 (0.84 - 0.97) | 0.007 |
| | 2021 | 13.3 | 1.25 (1.12 - 1.39) | <0.001 | 2.5 | 1.07 (0.84 - 1.35) | 0.629 | 5.4 | 1.38 (1.16 - 1.64) | <0.001 | 30.0 | 0.90 (0.83 - 0.97) | 0.004 |

**Supplementary Methods 1 – Interrupted Times Series Analysis**

The main analysis of this study compared incidence in 2021 to that in 2019, to obtain the risk ratio between a pre- COVID-19 year to one where the pandemic was ongoing. To understand to what extent the increase in risk ratio should be attributed to the pandemic, and to what extent it is part of an ongoing trend, the risk ratio between 2019 and 2017 was computed as reference.

The year 2020 was skipped, since it overlaps both the pre-pandemic period and the pandemic one. The first case of COVID-19 in Israel occurred 4 months into this period (recall that we start each year in Nov. 1$^{st}$ of the previous year). Moreover, initial lockdowns after the pandemic hit made health services less available. Hence, a reduction in diagnoses and prescriptions during the initial pandemic period may reflect a reduction access to health services rather than in mental distress.

The downside of this approach is that it is based on only three (albeit aggregated) values – incidence rates in 2017, 2019 and 2021. To analyze the data more fully we employ herein Interrupted Times Series (ITS) Analysis. Data points correspond to the beginning of each week, from Nov. 1$^{st}$ 2016 to Oct. 3$^{rd}$ 2021, and denote the incidence rate per 1,000 adolescents during the four weeks that follow. Two Fourier terms were included in the analysis to model seasonality and ordinary least-squares was used to fit the model to the data.

Importantly, this analysis depends on the modeling choices for the interruption and the changes that might follow – several alternatives are examined here:

1. The interruption is defined as occurring on Feb. 27$^{th}$ 2020, the day the first case of COVID-19 was detected in Israel. The model allows for both a level change and a trend change (figure S1). In this modeling, most resultant models fit the data with an initial negative level change, followed by a significant positive trend change. The exception to this is diagnoses of eating disorder, for which, even though there is a decrease in incidence during the lockdown period, the model yields a significant increase in both level and trend.
2. The interruption is defined as occurring on Feb. 27$^{th}$ 2020, but the data between that date and Apr. 30$^{th}$ (end of the first lockdown) is not taken to fit the model. This mitigates the reduction in diagnoses and prescriptions that is apparent during the lockdown, and which probably does not reflect a reduction in mental distress (figure

S2). In this modeling there is a negative level change in incidence of antipsychotic and ADHD medication and diagnoses, and a positive level change in the other measures. This change is then followed by an increase in trend, in all measures except eating disorders, where the trend decreases (not statistically significant) after an initial level change of more than 50%.

3. The interruption is defined as occurring on Feb. 27th 2020, and the data between Feb. 27th and May 31st 2020 is not taken to fit the model. This mitigates the potential effect of a “bounce” occurring in May, once health services became more readily available (figure S3). Resultant models are qualitatively similar to those in (2). The exceptions are the incidence of antipsychotic medication, which displays a positive level change in this case, and the incidence of depression diagnoses, which displays a negative change in trend (both not statistically significant).
4. The interruption and gap are defined as in (3). However, observing that for most diagnoses and prescriptions the incidence rate in June 2020 is similar to that in previous years we examine a model which allows only for trend change (figure S4). In this modeling, there is a positive and significant trend change in the incidence of all the examined medication and diagnoses. Note that this model is appropriate when the incidence during June 2020 is similar to what is expected by the trend of previous years. Namely, it is probably inappropriate for modeling the incidence of eating disorders, which increases well above previous years, but might be appropriate for the other measures.
5. The interruption and gap are defined as in (3). However, for some diagnoses and medications, such as those for ADHD, the periodicity of prescription and diagnoses incidence is strongly influenced by the school cycle. Namely, incidence is low during the summer break (July-August), and peaks in the winter months. School closures have apparently disrupted this cycle, and, accordingly, we examine a model where there are different coefficients for the Fourier terms, before and after the interruption (figure S5). This modeling suggests that the cycle of the ADHD incidence was indeed disrupted during the COVID-19 era, and, to a lesser extent, also in the prescription of antipsychotic medication.
6. The interruption is defined as occurring on Feb. 27th 2020, and a second interruption on March 7th 2021, the data between Feb. 27th and May 31st 2020 is not taken to fit

the model. As in (5), both level-change and trend-change are included after this interruption (figure S6). Vaccination efforts in Israel started relatively early, during the “third wave” of COVID-19 in December 2020[1]. By March 2021 it seemed that these efforts were very successfully. The third wave was curbed, and restrictions were mostly lifted. We examine a model where we allow a second interruption on March 7th, 2021, the day all schools resumed regular classes. We examined this setting without a level-change covariate, as we expected this effect to be gradual. We allow for one set of coefficients for the period before Feb 27th 2020 and after March 7th 2021, and a second set of coefficients for the intermediate period, May 31st 2020 – March 7th 2021 (trend-covariate is denoted as “vaccination trend” in eTable 2). In this modeling there is a significant positive level change for the incidence of depression, eating disorders, and ADHD diagnoses and medication. In addition, there is a significant increase in the trend for antidepressants, stress diagnoses, and ADHD diagnoses and medication. Interestingly, after the full reopening of schools on March 7th 2021, the trend decreases. Though this decrease is based on a relatively small number of data points, it is statistically significant for anxiety, stress and eating disorder diagnoses as well as antidepressants and antipsychotic medication and gives hope that the sharp increase in mental distress that is reflected in the data may be slowing down now that schools are once again open, and that the availability of vaccines gives us greater confidence in our ability to deal with the pandemic.

7. During May 2021 the Israeli-Palestinian conflict escalated into violent outbreaks throughout the country. The crisis started on the 6th of May, and is considered to be over by May 21st. Since this crisis may have further exacerbated the mental health situation is Israel, a final analysis in this venue is similar to (6), with the exception that data between May 6th and June 21st was excluded when fitting the model (allowing for an additional month in which the effect might be apparent). Results are nearly identical to (6) above, suggesting that the crisis did not have a direct effect on the underlying trend.

The parameters for these seven models are listed in the eTable 2.

**eFigure 1:** ITS analysis with an interruption on Feb. 27th, 2020, with level and trend change and no gap*

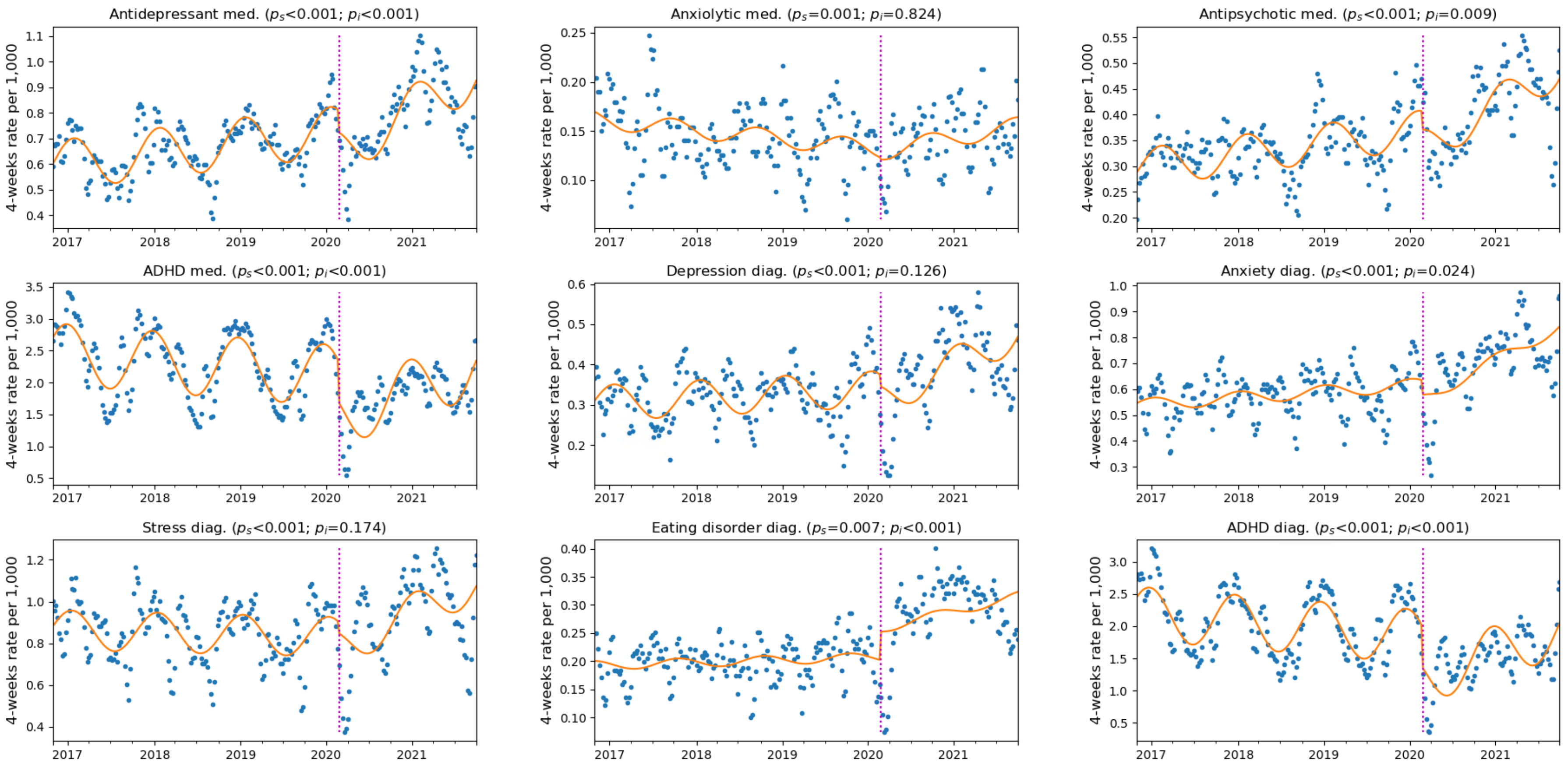


Legend: Interruption date indicated by a magenta dotted line. $P_s$ denotes the p-value for the change in trend, $P_i$ the p-value for change in level. Orange line depicts the fit of the ITS model.

* This figure is similar to Figure 3 in the main manuscript

**eFigure 2** - ITS analysis with an interruption on Feb. 27th, 2020, with gap in fitting the model between Feb. 27th and Apr. 30th, 2020

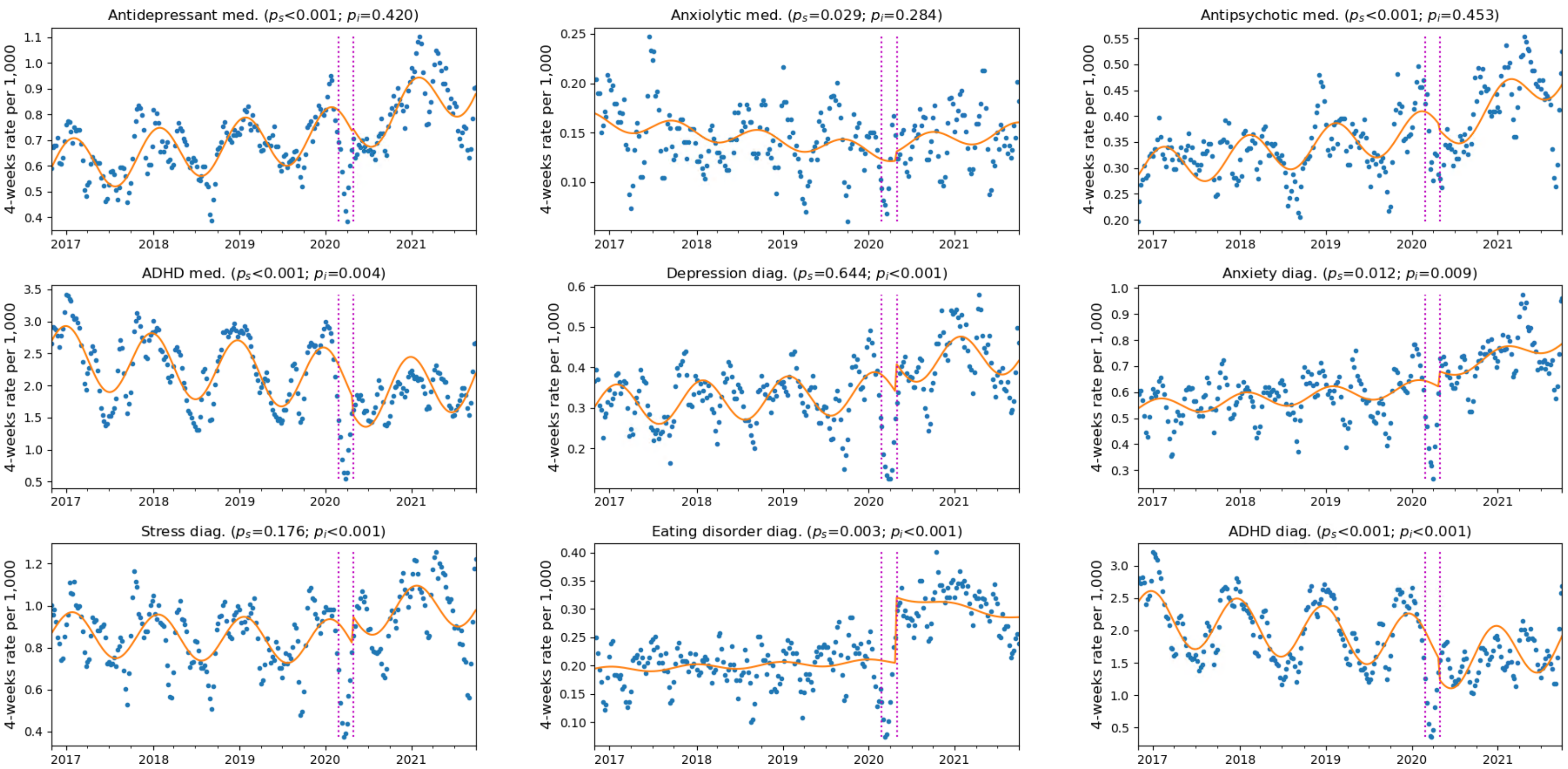


Legend: Left magenta line indicates the interruption date, after which data is ignored in modeling until the date corresponding to the right magenta line. $P_s$ denotes the p-value for the change in trend, $P_i$ the p-value for change in level. Orange line depicts the fit of the ITS model.

**eFigure 3** - ITS analysis with an interruption on Feb. 27th, 2020, with gap in fitting the model between Feb. 27th and May 31st, 2020

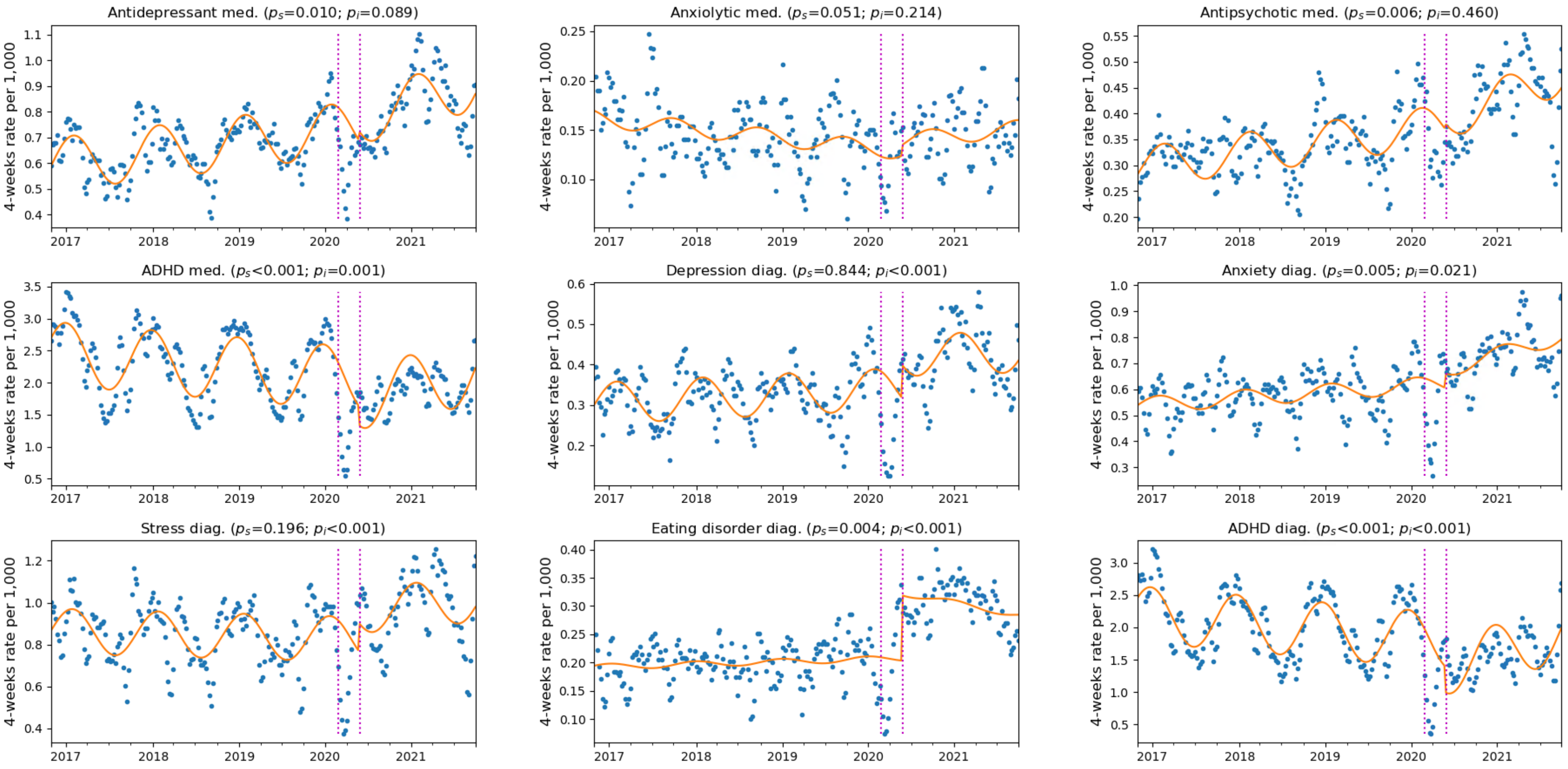


Legend: Left magenta line indicates the interruption date, after which data is ignored in modeling until the date corresponding to the right magenta line. $P_s$ denotes the p-value for the change in trend, $P_i$ the p-value for change in level. Orange line depicts the fit of the ITS model.

**eFigure 4** - ITS analysis with an interruption on Feb. 27th, 2020, with gap in fitting the model between Feb. 27th and May 31st, 2020, model does not include level-change at the interruption point

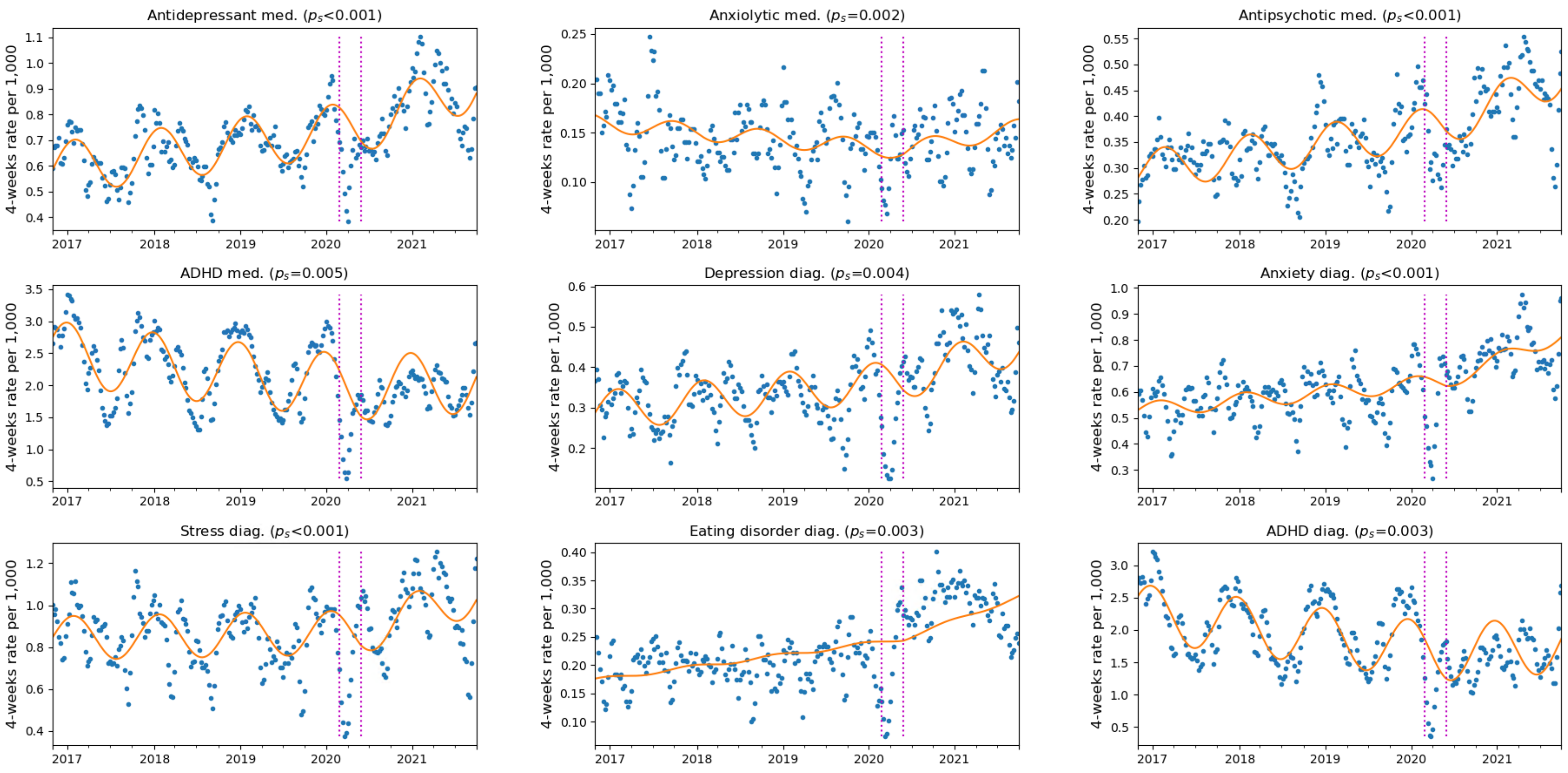


Legend: Left magenta line indicates the interruption date, after which data is ignored in modeling until the date corresponding to the right magenta line. $P_s$ denotes the p-value for the change in trend, $P_i$ the p-value for change in level. Orange line depicts the fit of the ITS model.

**eFigure 5** - ITS analysis with an interruption on Feb. 27th, 2020, with gap in fitting the model between Feb. 27th and May 31st, 2020, model includes different seasonal coefficients for the Fourier terms before and after the interruption point

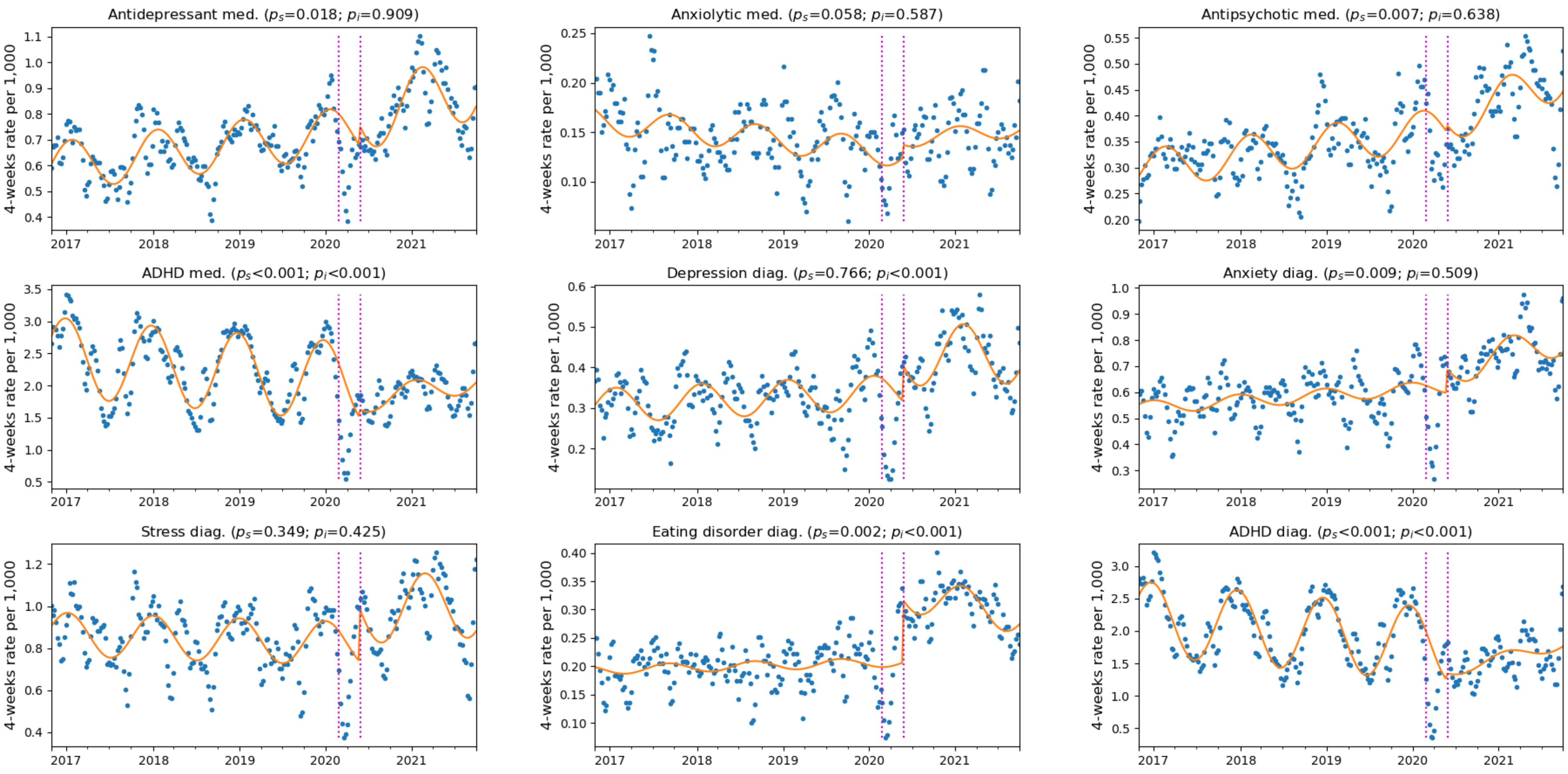


Legend: Left magenta line indicates the interruption date, after which data is ignored in modeling until the date corresponding to the right magenta line. $P_s$ denotes the p-value for the change in trend, $P_i$ the p-value for change in level. Orange line depicts the fit of the ITS model.

**eFigure 6** - ITS analysis with an interruption on Feb. 27th, 2020 and a second interruption on Mar. 7th 2021, with gap between Feb. 27th and May 31st 2020. Model includes level-change and trend-change at the first interruption point and only trend-change after the second interruption point. Model allows for different coefficients for the Fourier terms.

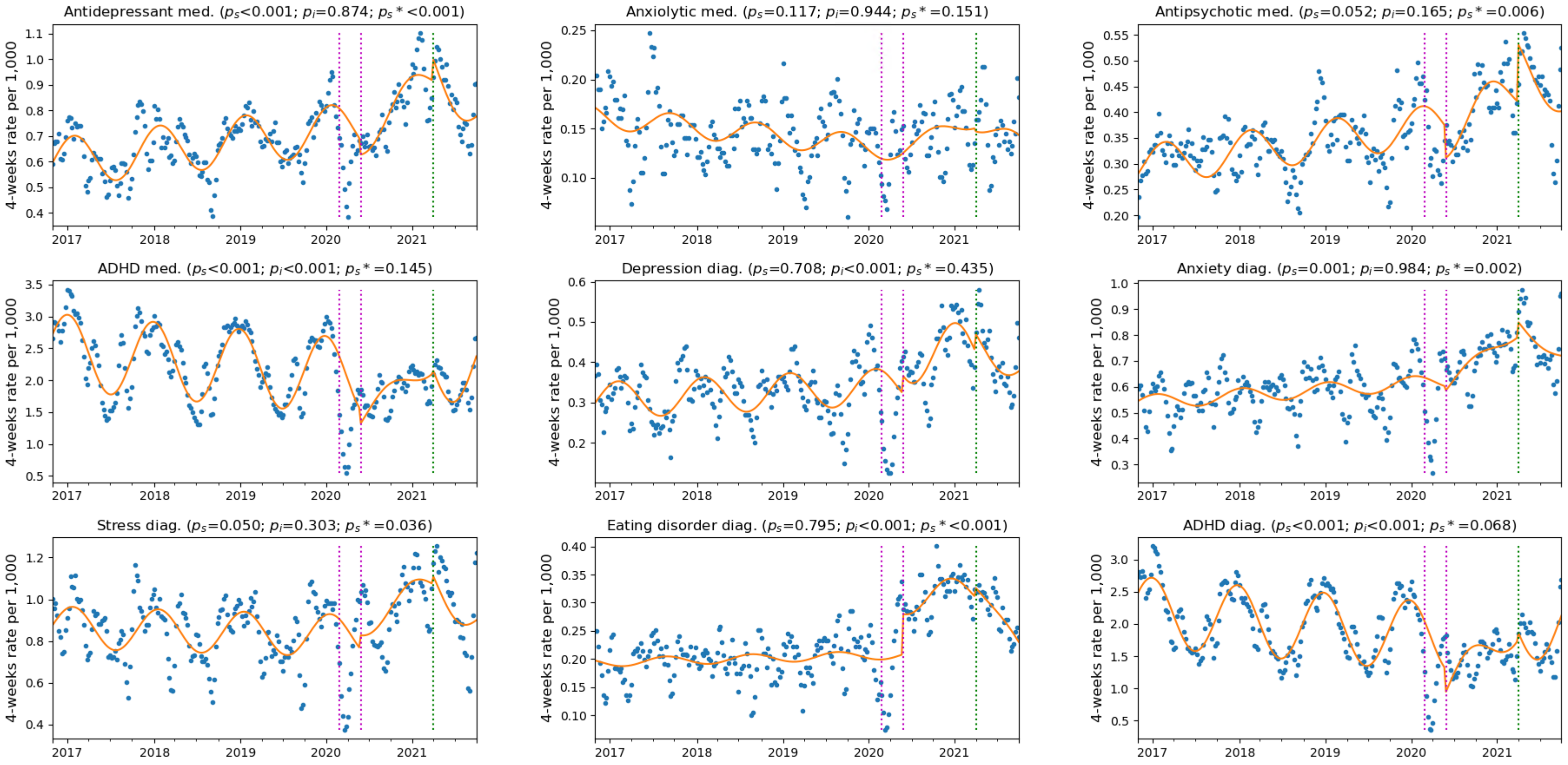


Legend: Left magenta line indicates the interruption date, after which data is ignored in modeling until the date corresponding to the right magenta line. Green line indicates the second interruption point. $P_s$ denotes the p-value for the change in trend after the first interruption, $P_i$ the p-value for change in level. $P_{s*}$ denotes the p-value for the change in trend after the second interruption. Orange line depicts the fit of the ITS model.

**eFigure 7** - ITS analysis with an interruption on Feb. 27th, 2020 and a second interruption on Mar. 7th 2021, with gap between Feb. 27th and May 31st 2020, and between May 6th 2021 and June 21st 2021. Model includes level-change and trend-change at the first interruption point and only trend-change after the second interruption point. Model allows for different coefficients for the Fourier terms.

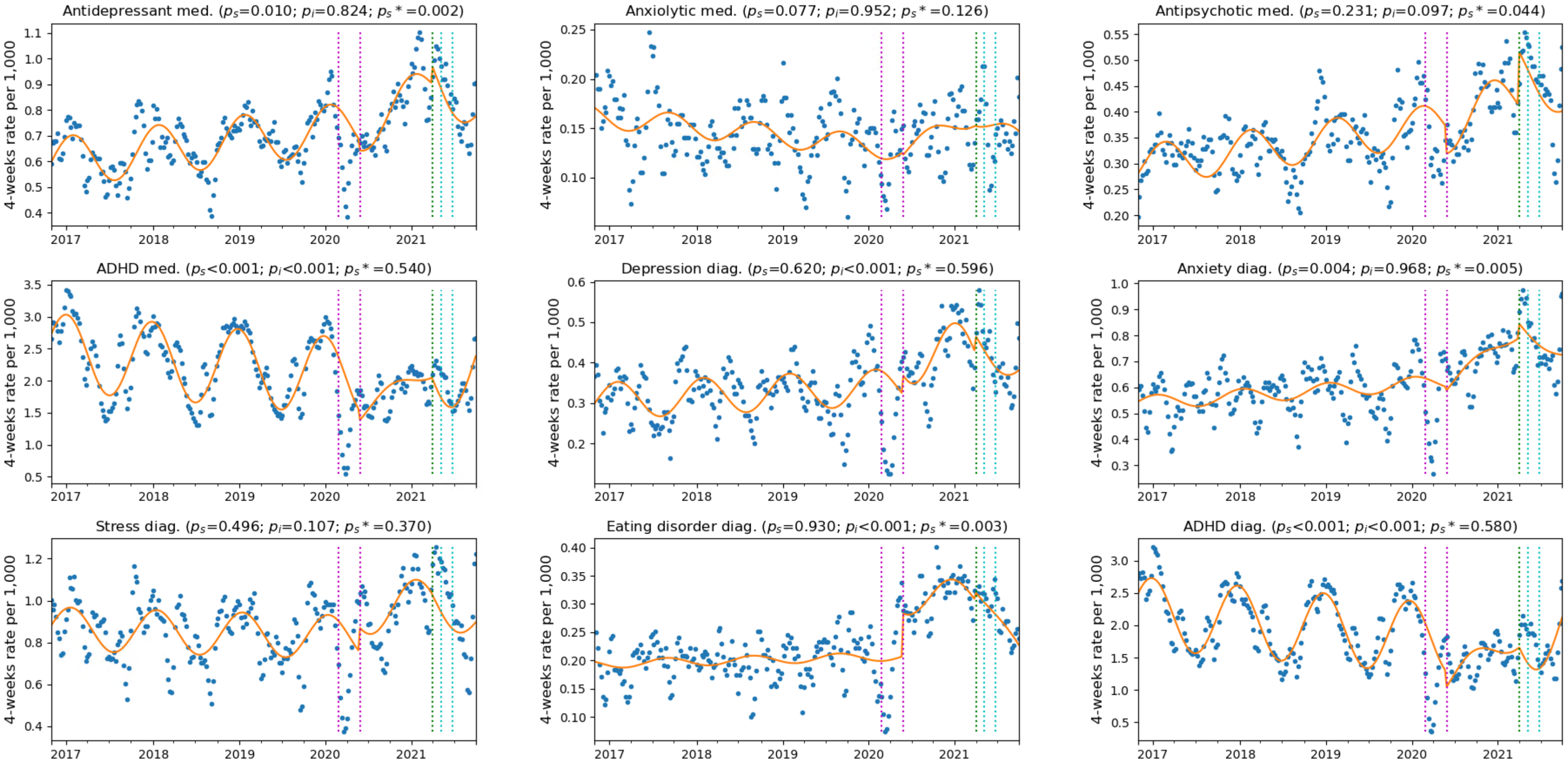


Legend: Left magenta line indicates the interruption date, after which data is ignored in modeling until the date corresponding to the right magenta line. Green line indicates the second interruption point. Cyan lines indicate the second gap period. $P_s$ denotes the p-value for the change in trend after the first interruption, $P_i$ the p-value for change in level. $P_{s*}$ denotes the p-value for the change in trend after the second interruption. Orange line depicts the fit of the ITS model.

**eTable 2:** Parameters for the ITS models with different modeling choices for data gap and interruption points

| | Antidepressants med | Anxiolytics med | Antipsychotics med | ADHD agents | Depression diag | Anxiety diag | Stress diag | Eating disorder diag | ADHD diag |
|---|---|---|---|---|---|---|---|---|---|
| **Model 1 - with interruption on Feb. 27$^{th}$, 2020, no gap** | | | | | | | | | |
| Prepandemic trend | 0.0008 (0.0005 - 0.0010) | -0.0002 (-0.0003 - -0.0001) | 0.0004 (0.0003 - 0.0006) | -0.0020 (-0.0030 - -0.0010) | 0.0002 (0.0000 - 0.0004) | 0.0005 (0.0002 - 0.0007) | -0.0002 (-0.0006 - 0.0002) | 0.0001 (-0.0000 - 0.0002) | -0.0021 (-0.0031 - -0.0011) |
| Pandemic level change | -0.0853 (-0.1293 - -0.0412) | -0.0018 (-0.0179 - 0.0143) | -0.0355 (-0.0622 - -0.0088) | -0.6330 (-0.8088 - -0.4573) | -0.0281 (-0.0641 - 0.0080) | -0.0555 (-0.1036 - -0.0074) | -0.0529 (-0.1293 - 0.0236) | 0.0509 (0.0267 - 0.0751) | -0.6333 (-0.8116 - -0.4551) |
| Pandemic trend change | 0.0030 (0.0022 - 0.0038) | 0.0005 (0.0002 - 0.0008) | 0.0014 (0.0010 - 0.0019) | 0.0115 (0.0084 - 0.0147) | 0.0018 (0.0012 - 0.0024) | 0.0029 (0.0021 - 0.0038) | 0.0040 (0.0026 - 0.0053) | 0.0006 (0.0002 - 0.0010) | 0.0111 (0.0079 - 0.0143) |
| **Model 2 - with interruption on Feb. 27th, 2020, with gap between Feb. 27th and Apr. 30th 2020** | | | | | | | | | |
| Prepandemic trend | 0.0008 (0.0005 - 0.0010) | -0.0002 (-0.0003 - -0.0001) | 0.0004 (0.0003 - 0.0006) | -0.0021 (-0.0031 - -0.0012) | 0.0002 (0.0000 - 0.0004) | 0.0005 (0.0002 - 0.0007) | -0.0002 (-0.0006 - 0.0002) | 0.0001 (-0.0000 - 0.0002) | -0.0022 (-0.0032 - -0.0013) |
| Pandemic level change | 0.0171 (-0.0246 - 0.0587) | 0.0091 (-0.0076 - 0.0259) | -0.0106 (-0.0383 - 0.0171) | -0.2590 (-0.4329 - -0.0851) | 0.0720 (0.0401 - 0.1038) | 0.0604 (0.0153 - 0.1055) | 0.1346 (0.0638 - 0.2053) | 0.1161 (0.0959 - 0.1362) | -0.3075 (-0.4870 - -0.1280) |
| Pandemic trend change | 0.0015 (0.0007 - 0.0023) | 0.0004 (0.0000 - 0.0007) | 0.0012 (0.0006 - 0.0017) | 0.0065 (0.0031 - 0.0099) | 0.0001 (-0.0005 - 0.0008) | 0.0011 (0.0003 - 0.0020) | 0.0010 (-0.0004 - 0.0023) | -0.0006 (-0.0010 - -0.0002) | 0.0069 (0.0034 - 0.0104) |
| **Model 3 - with interruption on Feb. 27th, 2020, with gap between Feb. 27th and May 31st 2020** | | | | | | | | | |
| Prepandemic trend | 0.0008 (0.0006 - 0.0010) | -0.0002 (-0.0003 - -0.0001) | 0.0004 (0.0003 - 0.0006) | -0.0022 (-0.0031 - -0.0012) | 0.0002 (0.0000 - 0.0004) | 0.0004 (0.0002 - 0.0007) | -0.0002 (-0.0006 - 0.0002) | 0.0001 (-0.0000 - 0.0002) | -0.0022 (-0.0032 - -0.0013) |
| Pandemic level change | 0.0371 (-0.0057 - 0.0799) | 0.0110 (-0.0064 - 0.0283) | 0.0105 (-0.0175 - 0.0385) | -0.2936 (-0.4721 - -0.1151) | 0.0821 (0.0495 - 0.1148) | 0.0548 (0.0083 - 0.1013) | 0.1366 (0.0635 - 0.2096) | 0.1151 (0.0943 - 0.1359) | -0.3972 (-0.5780 - -0.2164) |
| Pandemic trend change | 0.0012 (0.0003 - 0.0021) | 0.0004 (-0.0000 - 0.0007) | 0.0008 (0.0002 - 0.0014) | 0.0079 (0.0042 - 0.0116) | -0.0001 (-0.0007 - 0.0006) | 0.0014 (0.0004 - 0.0023) | 0.0010 (-0.0005 - 0.0025) | -0.0006 (-0.0011 - -0.0002) | 0.0095 (0.0058 - 0.0133) |
| **Model 4 - with interruption on Feb. 27th, 2020, with gap between Feb. 27th and May 31st 2020, no level-change at the** | | | | | | | | | |
| Prepandemic trend | 0.0009 (0.0007 - 0.0011) | -0.0002 (-0.0002 - -0.0001) | 0.0005 (0.0003 - 0.0006) | -0.0029 (-0.0037 - -0.0021) | 0.0004 (0.0003 - 0.0006) | 0.0006 (0.0004 - 0.0008) | 0.0002 (-0.0002 - 0.0005) | 0.0004 (0.0003 - 0.0005) | -0.0033 (-0.0041 - -0.0025) |
| Pandemic trend change | 0.0016 (0.0008 - 0.0023) | 0.0005 (0.0002 - 0.0008) | 0.0009 (0.0004 - 0.0014) | 0.0045 (0.0014 - 0.0077) | 0.0009 (0.0003 - 0.0015) | 0.0020 (0.0012 - 0.0028) | 0.0025 (0.0013 - 0.0038) | 0.0007 (0.0002 - 0.0011) | 0.0050 (0.0017 - 0.0082) |
| **Model 5 - with interruption on Feb. 27th, 2020, with gap between Feb. 27th and May 31st 2020, different Fourier coefficients** | | | | | | | | | |
| Prepandemic trend | 0.0008 (0.0005 - 0.0010) | -0.0002 (-0.0003 - -0.0001) | 0.0004 (0.0003 - 0.0006) | -0.0022 (-0.0030 - -0.0013) | 0.0002 (0.0000 - 0.0004) | 0.0004 (0.0002 - 0.0007) | -0.0003 (-0.0006 - 0.0001) | 0.0001 (-0.0000 - 0.0002) | -0.0023 (-0.0031 - -0.0015) |
| Pandemic level change | -0.0028 (-0.0518 - 0.0461) | 0.0055 (-0.0145 - 0.0255) | 0.0079 (-0.0251 - 0.0408) | -0.5743 (-0.7615 - -0.3872) | 0.0698 (0.0320 - 0.1075) | 0.0179 (-0.0356 - 0.0714) | 0.0331 (-0.0485 - 0.1147) | 0.1119 (0.0889 - 0.1349) | -0.7402 (-0.9235 - -0.5569) |
| Pandemic trend change | 0.0010 (0.0002 - 0.0019) | 0.0003 (-0.0000 - 0.0007) | 0.0008 (0.0002 - 0.0014) | 0.0070 (0.0036 - 0.0103) | -0.0001 (-0.0008 - 0.0006) | 0.0013 (0.0003 - 0.0022) | 0.0007 (-0.0008 - 0.0021) | -0.0006 (-0.0010 - -0.0002) | 0.0084 (0.0052 - 0.0117) |
| **Model 6 - with interruption on Feb. 27$^{th}$, 2020 and on Mar. 7$^{th}$ 2021, with gap between Feb. 27$^{th}$ and May 31$^{st}$ 2020, no level-change at the second interruption point, different Fourier coefficients** | | | | | | | | | |
| Prepandemic trend | 0.0008 (0.0006 - 0.0010) | -0.0002 (-0.0003 - -0.0001) | 0.0004 (0.0003 - 0.0006) | -0.0021 (-0.0030 - -0.0013) | 0.0002 (0.0000 - 0.0004) | 0.0004 (0.0002 - 0.0007) | -0.0002 (-0.0006 - 0.0002) | 0.0001 (-0.0000 - 0.0002) | -0.0022 (-0.0031 - -0.0014) |
| Pandemic level change | -0.0064 (-0.0854 - 0.0726) | 0.0012 (-0.0321 - 0.0345) | 0.0357 (-0.0148 - 0.0862) | -0.7521(-1.0657 - -0.4386) | 0.1140 (0.0517 - 0.1762) | -0.0009 (-0.0890 - 0.0872) | 0.0736 (-0.0669 - 0.2141) | 0.1141 (0.0774 - 0.1509) | -0.8740 (-1.1868 - -0.5613) |
| Pandemic trend change | 0.0045 (0.0020 - 0.0070) | 0.0008 (-0.0002 - 0.0019) | 0.0016 (-0.0000 - 0.0032) | 0.0226 (0.0127 - 0.0324) | -0.0004 (-0.0023 - 0.0016) | 0.0046 (0.0019 - 0.0074) | 0.0044 (-0.0000 - 0.0088) | 0.0002 (-0.0010 - 0.0013) | 0.0258 (0.0159 - 0.0356) |
| Vaccination trend change | -0.0111 (-0.0164 - -0.0058) | -0.0016 (-0.0039 - 0.0006) | -0.0048 (-0.0082 - -0.0014) | -0.0157 (-0.0368 - 0.0054) | -0.0017 (-0.0059 - 0.0025) | -0.0097 (-0.0156 - -0.0037) | -0.0101 (-0.0196 - -0.0006) | -0.0044 (-0.0068 - -0.0019) | -0.0196 (-0.0407 - 0.0014) |
| **Model 7 - with interruption on Feb. 27$^{th}$, 2020 and on Mar. 7$^{th}$ 2021, with gap between Feb. 27$^{th}$ and May 31$^{st}$ 2020, and between May 6th and June 21st 2021. No level-change at the second interruption point, different Fourier coefficients** | | | | | | | | | |
| Prepandemic trend | 0.0008 (0.0006 - 0.0010) | -0.0002 (-0.0003 - -0.0001) | 0.0004 (0.0003 - 0.0006) | -0.0021 (-0.0030 - -0.0013) | 0.0002 (0.0000 - 0.0004) | 0.0004 (0.0002 - 0.0007) | -0.0002 (-0.0006 - 0.0001) | 0.0001 (-0.0000 - 0.0002) | -0.0022 (-0.0031 - -0.0014) |
| Pandemic level change | 0.0091 (-0.0715 - 0.0897) | -0.0010 (-0.0343 - 0.0323) | 0.0438 (-0.0081 - 0.0956) | -0.6885 (-1.0085 - -0.3686) | 0.1165 (0.0527 - 0.1803) | 0.0019 (-0.0888 - 0.0926) | 0.1166 (-0.0253 - 0.2585) | 0.1172 (0.0797 - 0.1548) | -0.7721 (-1.0857 - -0.4585) |
| Pandemic trend change | 0.0035 (0.0008 - 0.0062) | 0.0010 (-0.0001 - 0.0021) | 0.0010 (-0.0007 - 0.0028) | 0.0185 (0.0079 - 0.0290) | -0.0005 (-0.0026 - 0.0016) | 0.0045 (0.0015 - 0.0075) | 0.0016 (-0.0031 - 0.0063) | -0.0001 (-0.0013 - 0.0012) | 0.0191 (0.0088 - 0.0295) |
| Vaccination trend change | -0.0091 (-0.0147 - -0.0034) | -0.0018 (-0.0042 - 0.0005) | -0.0037 (-0.0074 - -0.0001) | -0.0070 (-0.0295 - 0.0155) | -0.0012 (-0.0057 - 0.0033) | -0.0091 (-0.0155 - -0.0028) | -0.0045 (-0.0145 - 0.0054) | -0.0041 (-0.0067 - -0.0014) | -0.0062 (-0.0282 - 0.0158) |

**Supplementary Methods 2 -** Distributions of new diagnoses and prescriptions according to physician's profession

Mental health diagnoses and prescriptions can be given not only by psychiatrists, but also by doctors with other types of specializations, such as general practitioners or pediatricians. eFigure 7 depicts the distribution of new diagnoses and prescriptions according to the physician's profession. This data is not always listed, and cases where it was missing were discarded. It is important to note that some patients may be initially diagnosed in a private practice, and in such cases the entry that we see in the EHR may actually be a reaffirmation of a diagnoses, rather than a new one.

**eFigure 8 -** Distribution of new diagnoses and prescriptions according to physician's profession, during the periods of November 1, 2016 - October 31, 2017 (a), November 1, 2018 -October 31, 2019 (b), November 1, 2020 – October 31, 2021 (c).

a. Period November 1, 2016 -October 31, 2017

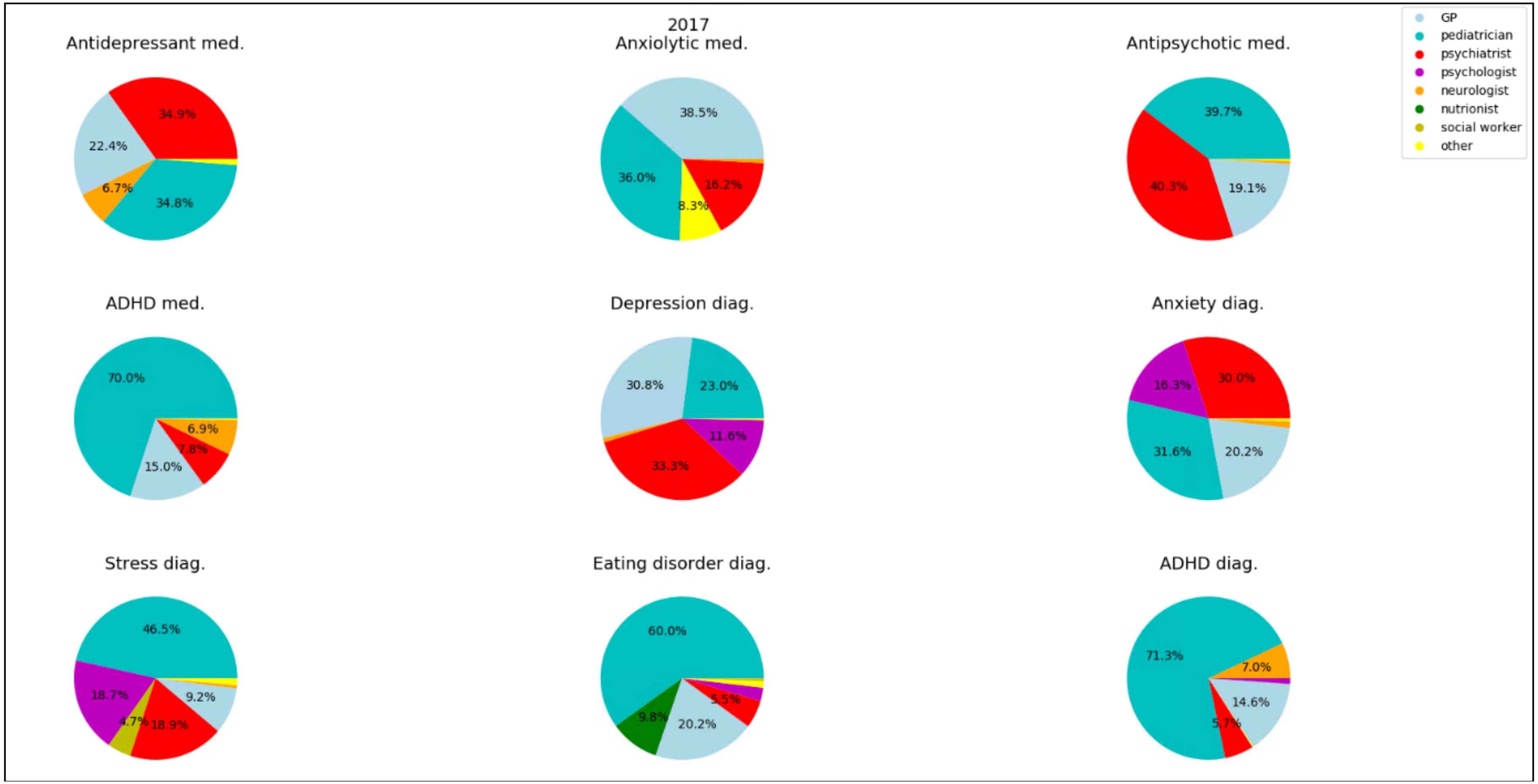

b. Period November 1, 2018 -October 31, 2019

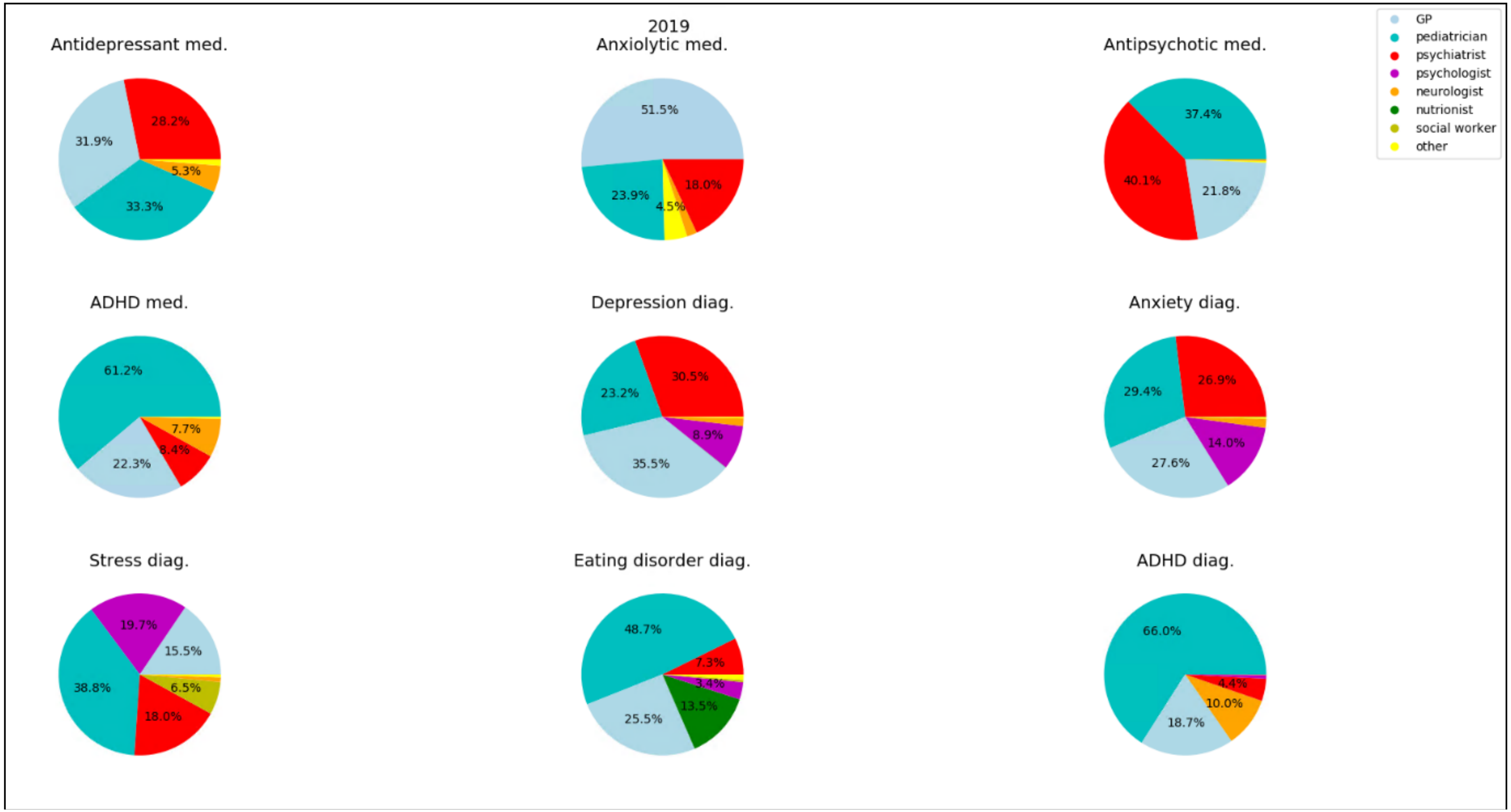

c. Period November 1, 2020 – October 31, 2021

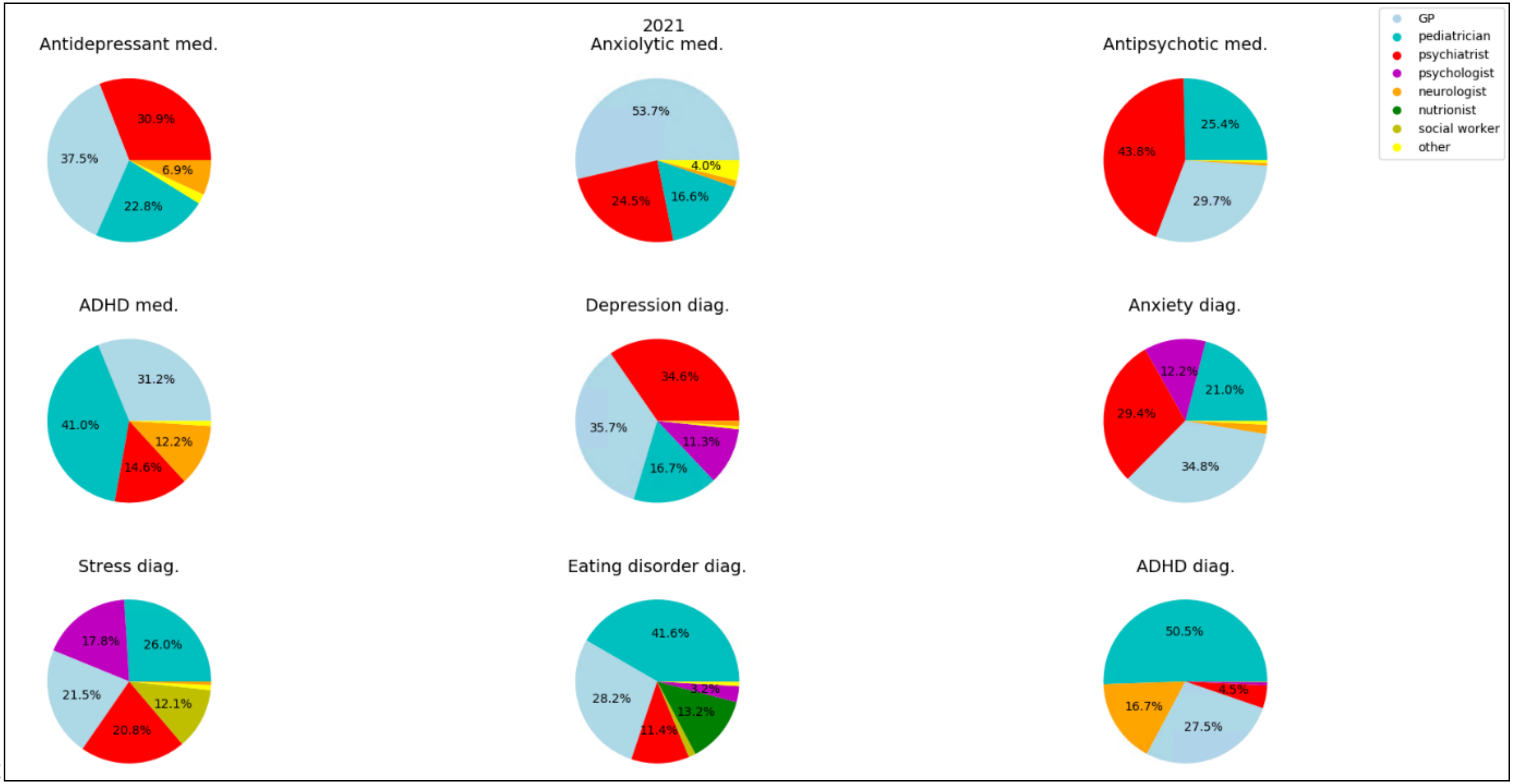

C

**eTable3** COVID-19 Israel Policy Restrictions

| Interval | Date begins | Date ends | Restrictions |
|---|---|---|---|
| Lockdown 1 | March 14, 2020 | May 04, 2020 | All schools closed, workplaces closed, gatherings banned, out-of-home distance restrictions, international travel banned |
| Lockdown 2 | September 18, 2020 | October 17, 2020 | |
| Lockdown 3 | December 27, 2020 | February 07, 2021 | |

All schools closed except nurseries. Workplaces closed for all but essential workplaces[2]
